# Untargeted metabolomics of COVID-19 patient serum reveals potential prognostic markers of both severity and outcome

**DOI:** 10.1101/2020.12.09.20246389

**Authors:** Ivayla Roberts, Marina Wright Muelas, Joseph M. Taylor, Andrew S. Davison, Yun Xu, Justine M. Grixti, Nigel Gotts, Anatolii Sorokin, Royston Goodacre, Douglas B. Kell

## Abstract

The diagnosis of COVID-19 is normally based on the qualitative detection of viral nucleic acid sequences. Properties of the host response are not measured but are key in determining outcome. Although metabolic profiles are well suited to capture host state, most metabolomics studies are either underpowered, measure only a restricted subset of metabolites, compare infected individuals against uninfected control cohorts that are not suitably matched, or do not provide a compact predictive model.

Here we provide a well-powered, untargeted metabolomics assessment of 120 COVID-19 patient samples acquired at hospital admission. The study aims to predict the patient’s infection severity (i.e., mild or severe) and potential outcome (i.e., discharged or deceased).

High resolution untargeted LC-MS/MS analysis was performed on patient serum using both positive and negative ionization modes. A subset of 20 intermediary metabolites predictive of severity or outcome were selected based on univariate statistical significance and a multiple predictor Bayesian logistic regression model was created. The predictors were selected for their relevant biological function and include cytosine and ureidopropionate (indirectly reflecting viral load), kynurenine (reflecting host inflammatory response), and multiple short chain acylcarnitines (energy metabolism) among others.

Currently, this approach predicts outcome and severity with a Monte Carlo cross validated area under the ROC curve of 0.792 (SD 0.09) and 0.793 (SD 0.08), respectively. A blind validation study on an additional 90 patients predicted outcome and severity at ROC AUC of 0.83 (CI 0.74 – 0.91) and 0.76 (CI 0.67 – 0.86). Prognostic tests based on the markers discussed in this paper could allow improvement in the planning of COVID-19 patient treatment.

## Introduction

The severe acute respiratory syndrome coronavirus-2 (SARS-CoV-2) outbreak started in Wuhan, China, in 2019, and quickly resulted in a worldwide pandemic, challenging healthcare systems with the need to provide intensive care to a previously inconceivable number of patients (Bennet et al., 2020). SARS-CoV-2 presents with a wide range of symptoms. These range from minor, unspecific ones, including anosmia, a dry persistent cough, fever, diarrhoea, in certain cases combined with mild pneumonia, to more severe, potentially life-threatening symptoms, such as severe pneumonia with dyspnoea, tachypnoea and disturbed gas exchange. Approximately 5% of severely infected patients develop lung dysfunction, requiring ventilation, and shock or multiple organ failure (Marietta et al., 2020; Wu and McGoogan, 2020). In some cases, symptoms remain for an extended period (‘long COVID’).

The reasons behind the wide variability in individual responses to COVID-19, i.e., the illness resulting from SARS-CoV-2 infection, are still poorly understood, though some appear to involve interferon responses (Arunachalam et al., 2020; Hadjadj et al., 2020; Zhang et al., 2020). Much research, and evidence from the clinic, points towards the idea that severe complications in COVID-19 arise through a vasculopathy and coagulopathy elicited by infection rather than via the typical inflammatory responses normally observed in acute respiratory distress syndrome or cytokine release storms (Fox et al., 2020; Grobler et al., 2020; Leisman et al., 2020; Libby and Lüscher, 2020; Paranjpe et al., 2020; Pretorius et al., 2020; Zheng et al., 2020). Prognostic scores attempt to transform complex clinical pictures into tangible numerical values. However, many of these novel COVID-19 prognostic scores have been found to have a high risk of bias, possibly reflecting the fact that they have been developed in small cohorts, and many have been published without clear details of model derivation and testing (Knight et al., 2020; Wynants et al., 2020).

Understanding changes in the biochemistry of an individual who is ostensibly healthy (Dunn et al., 2011; Dunn et al., 2015), including when they may show no overt symptoms of infection with SARS-CoV-2, remains a huge challenge (Alene et al., 2021). Similar questions apply to understanding who is likely to survive (unaided or via intervention) and who is likely to die from COVID-19 once diagnosed.

For fundamental reasons, the metabolome is a more sensitive indicator of the biochemical status of a cell or organism than is a proteome or a transcriptome (Kell and Oliver, 2016; Oliver et al., 1998; Raamsdonk et al., 2001). Consequently, metabolomic analyses of patient samples promise to enable understanding of biochemical changes in relation to poorly understood processes, for instance as recently shown in human frailty in ageing populations (Rattray et al., 2019). Metabolomics studies measure the effects on the host and not simply the presence of the infecting agent, therefore, such studies could provide a set of markers that can be of significant use for rapid tests, complementary to current polymerase chain reaction (PCR) or antibody tests, for confirmation of SARS-CoV-2 infection, disease severity, and potential outcome.

A continuously increasing number of studies have applied metabolomics to investigate COVID-19 in human patients; a selection of which is presented in Table 1. Most of the studies have highlighted disruption of lipid metabolism (López-Hernández et al., 2021; Overmyer et al., 2020; Thomas et al., 2020), along with tryptophan metabolism in relation to inflammation (Ansone et al., 2021; Blasco et al., 2020; López-Hernández et al., 2021; Overmyer et al., 2020; Sindelar et al., 2021; Thomas et al., 2020) and changes in pyrimidine metabolism (Blasco et al., 2020) as metabolic features of COVID-19 patients against controls. Some of these studies were clearly limited by patient numbers and may therefore not be fully representative of the variation in human responses to COVID-19, and some lacked proper statistical tests see (Broadhurst and Kell, 2006). Moreover, many of these markers of COVID-19 infection, severity and outcome have also been described in patients with sepsis and acute respiratory failure (Migaud et al., 2020). Most importantly, many of the early studies simply compare patients with healthy controls, which, given that the disease status is in fact known (and current diagnostics very rapid), does not of itself have either diagnostic or prognostic value. However, more recent studies with larger cohorts such as (López-Hernández et al., 2021; Sindelar et al., 2021) have also considered more interesting aspects such as severity and included longitudinal follow up of the infection. Since presence of the disease is already known, our work here is focused on this more pertinent question: can the metabolome distinguish patients with COVID-19 in terms of either disease severity (mild/severe) or outcome (deceased/survived)?

**Table 1.**
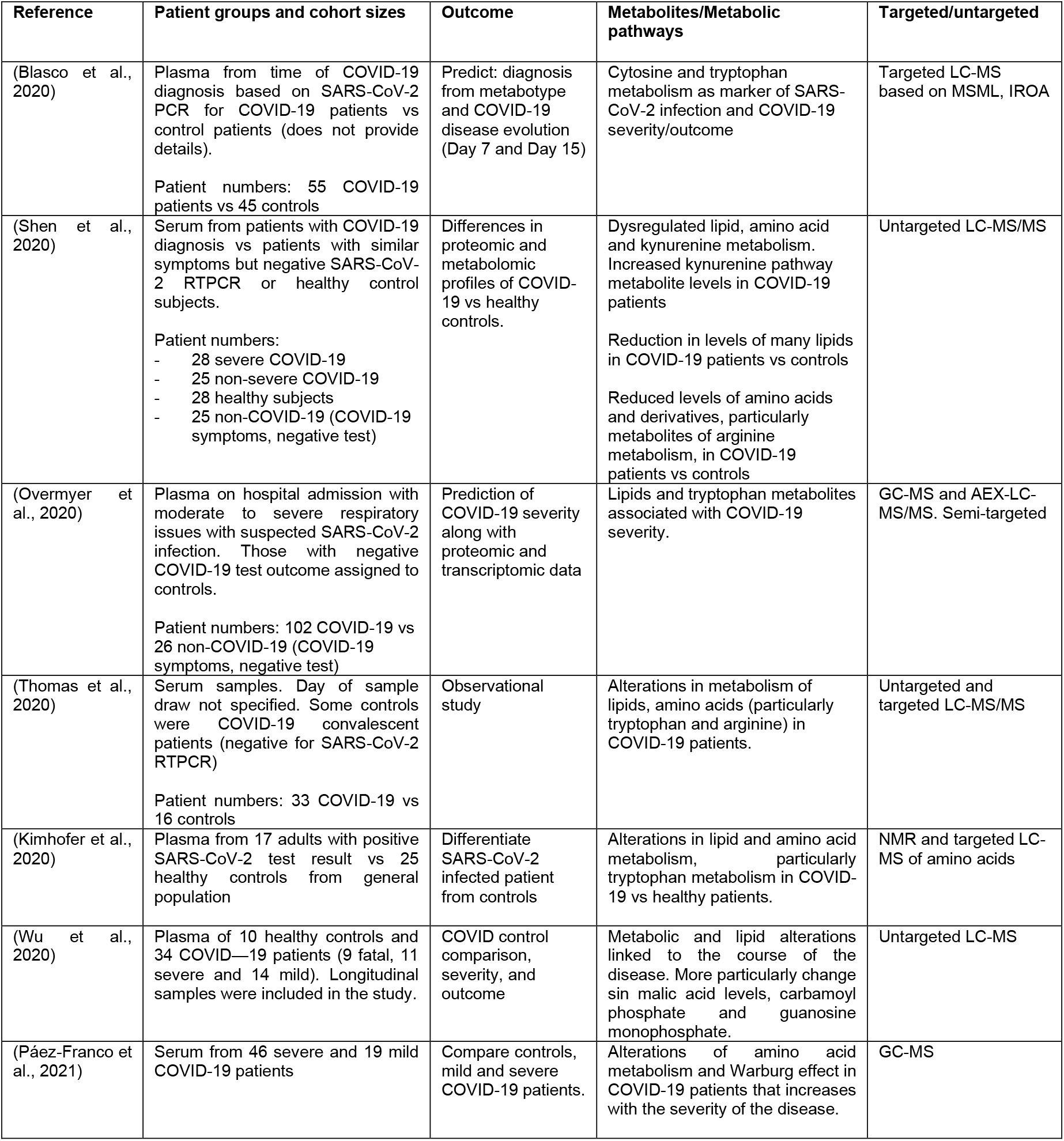
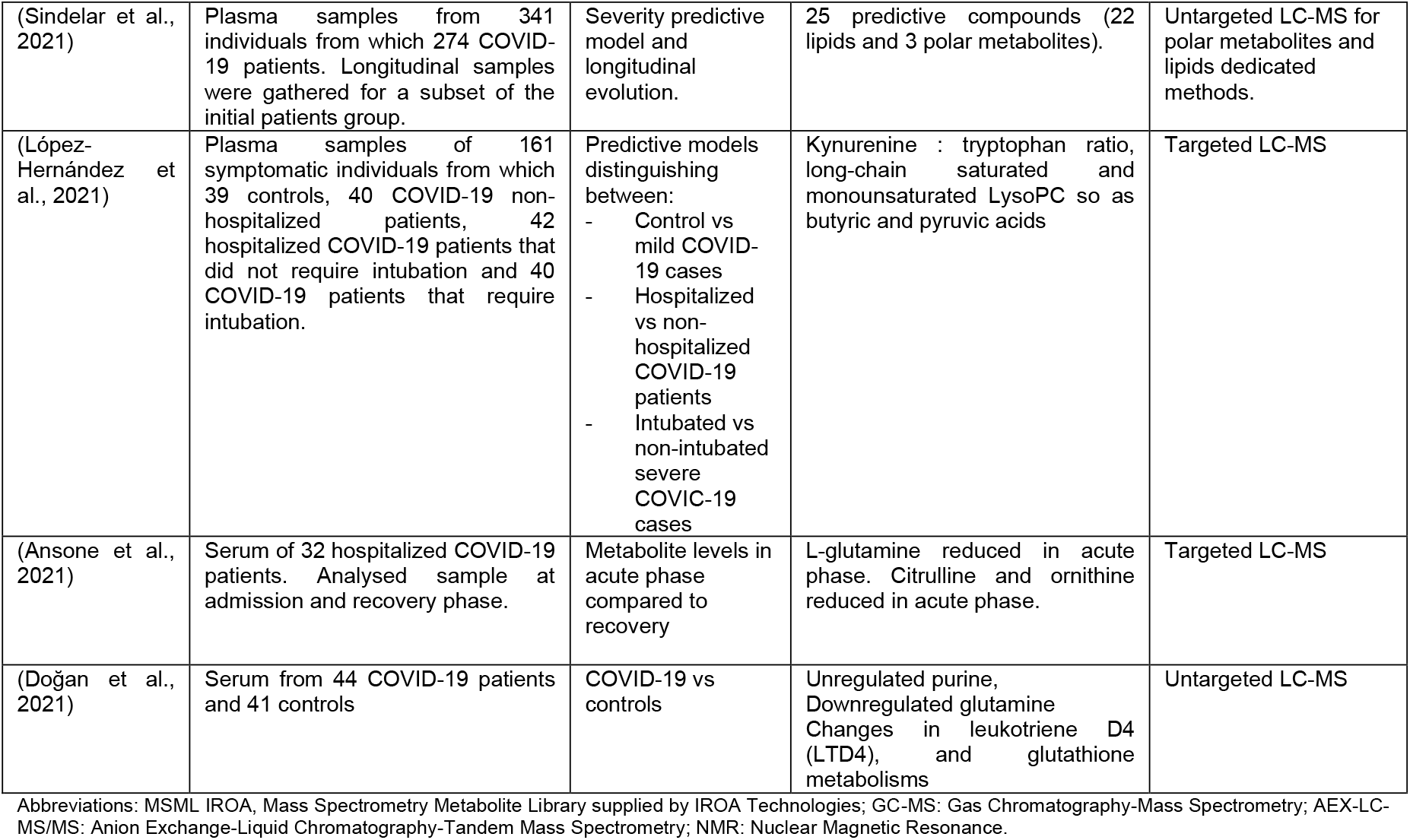
Overview of findings and study design in published studies utilising metabolomics to assess COVID-19 diagnosis, severity and outcome.

To that end, we adopted an untargeted metabolomics approach using LC-MS/MS to cohorts of serum samples collected at the Royal Liverpool University Hospital (RLUH). 120 patient samples were obtained at the time of admission and diagnosis with COVID-19. This study enabled us to identify prognostic biomarkers of both COVID-19 severity (severe *vs* mild) and outcome. Subsequently, the findings were validated in a blind study on 90 additional patients. Moreover, longitudinal data were acquired from 28 severe patients with divergent outcome, i.e., 13 deceased and 15 discharged to explore further the temporal evolution of the selected predictors.

Here we present the study results via a set of different models. We first validate that untargeted metabolomics provides better severity and outcome distinction *vs* using solely demographics and clinical data, via multiblock chemometric analysis. A high dimensional predictive model is then presented, based on more than 900 metabolites, showing promising predictive performance. Next, with the goal of clinical application in mind, we present predictive model results of 20 compounds selected largely on the basis of our confidence in the metabolites’ identity and known biological function. We discuss in detail the approaches used to make this selection via pathway enrichment analysis and manual curation. Moreover, we explore (and largely discount) the possible confounding effects of demographic factors and underlying conditions of the selected metabolic predictors. Finally, we validate the predictive power of the selected compounds in a blind study of a new patient cohort.

## Results

### Discovery study

Pre-processing of LC-MS data in Compound Discoverer resulted in 5234 positive electrospray ionization (ESI+) and 2465 negative electrospray ionization (ESI-) retained metabolic features. The excluded features resulting from the standard filters; i.e., background compounds, features with quality control (QC) samples’ coefficient of variation (CV) >30%, or its presence in less than 80% of the QCs. Additionally, compounds detected in less than 25% of the experimental samples were also excluded. The latter allowed the removal of a large number of metabolic features related to drugs and their metabolites taken by the patients for pre-existing conditions, which would otherwise simply have represented confounders.

### Exploratory analysis

Principal components analysis (PCA) was performed on both ESI+ and ESI-data and no clear clustering due to either severity or outcome could be observed (Figure S1), indicating that a simple, unsupervised method such as PCA had failed to detect differences in metabolic profiles related to COVID-19 infection and potential outcome in complex data resulting from LC-MS analysis. In contrast, when PCA was applied to the metadata associated with the 120 patients which include gender, age, body mass index (BMI), pulse, temperature, blood pressure, respiratory rate (full list is provided in the methods section), a trend of separation between severe and mild patients was observed (Figure 1A). This alone is not overly interesting because the severity assessment was drawn from a few variables in these metadata. However, when the metadata and the LC-MS data were analyzed together by using a multiblock PCA (Smilde et al., 2005; Xu and Goodacre, 2012) the separation trend improved (Figure 1B), indicating that there is relevant discriminatory information within the LC-MS data which could not be revealed by PCA had been discovered in a multiblock model with presence of a meta data block. Those observations are also valid in fatal outcomes where no clear clustering is observed in the metadata PCA block (Figure 1A). Individual block figures from the multiblock analysis are available in the supplementary information (Figure S2). Clearly PCA is not a classification technique and the lack or presence of clusters does not translate directly to any predictive potential of the data. To explore if differences in metabolic profiles are capable of predicting the severity and outcome of COVID-19 infection, the next section explores in detail predictive models based on the LC-MS data.

**Figure 1.**
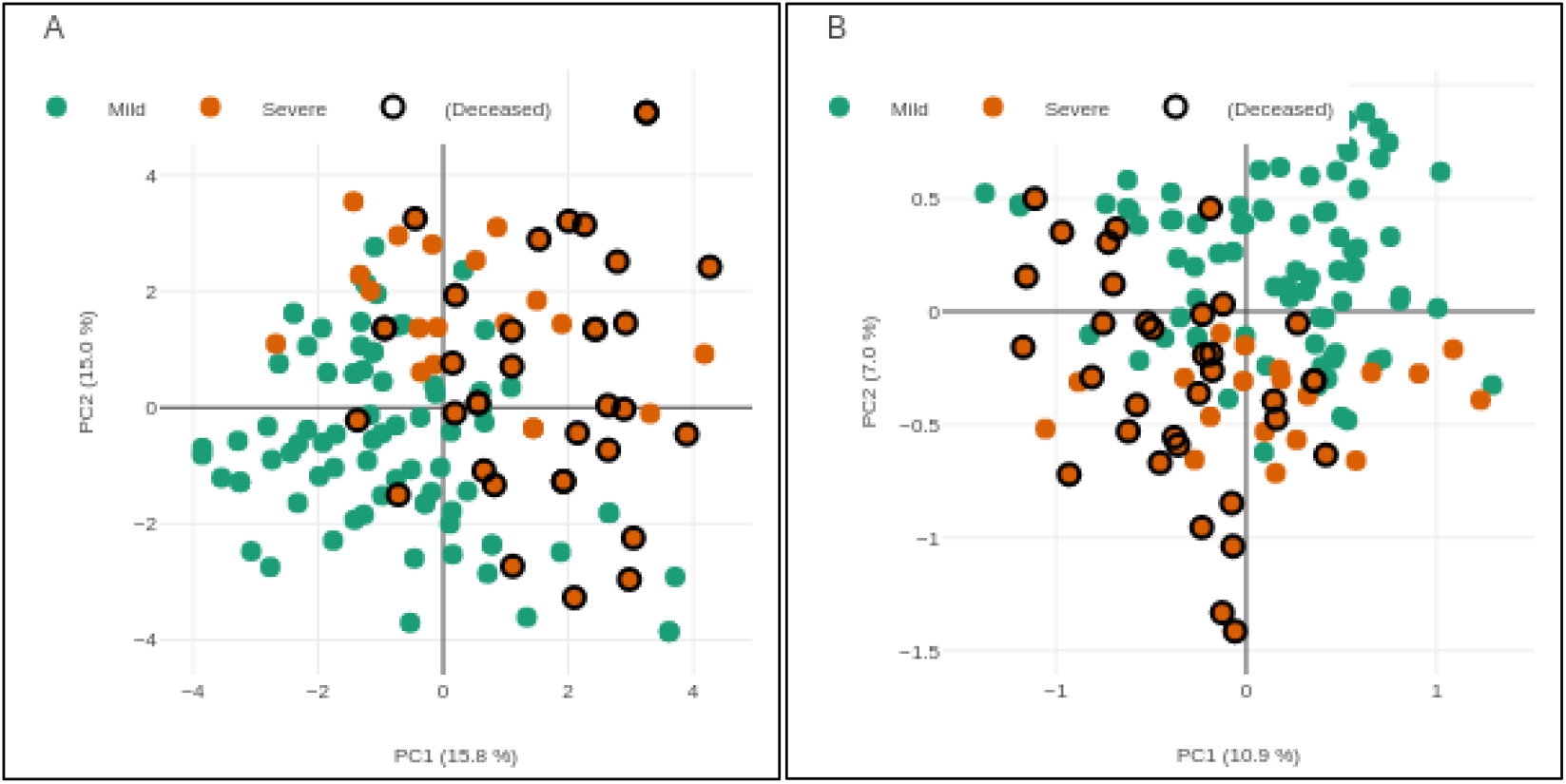
Metadata alone (A) and multiblock (B) PCA for severity and outcome. It can be observed that group separation trends improve with addition of LC-MS data (B). It is important to note that deceased patients are a subset of severe cases that do not appear to show specific clustering in this exploratory analysis. Axis are labelled with principal component and its explained variance in percentage.

### Predictive models

Four multi-predictor models were trained: extreme gradient boosted tree (Chen and Guestrin, 2016; Friedman, 2001), Lasso regularization with elastic net (Zou and Hastie, 2005), logistic regression, and Bayesian logistic regression (Goodrich, 2020). All these models, when trained on the complete data of >7000 metabolic features, showed clear signs of overfitting despite the use of cross-validation and regularization. To overcome this, the set of predictors was filtered based on individual significance as determined by volcano plot analysis (p-value and fold change) as illustrated in Figure 2 for outcome. Volcano plots for severity are available in the Supplementary information (Figure S3).

**Figure 2.**
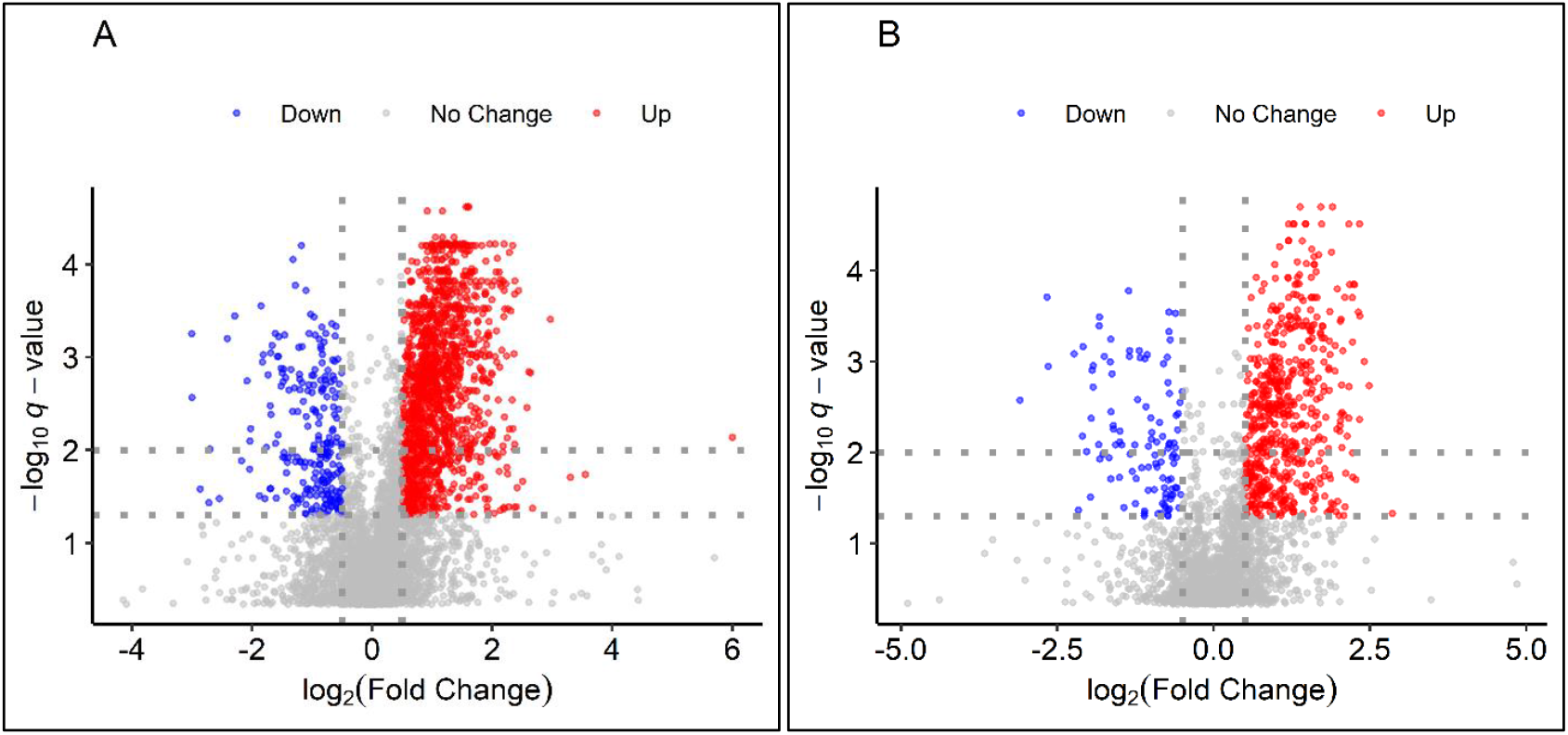
Volcano plots ESI+(A) and ESI-(B) for metabolites discriminating disease outcome. Dotted lines mark boundary of significance 0.5 for Log_2_ Fold Change (FC) and 2 lines for q-value at 0.05 and 0.01. Compounds with higher levels in poor outcome are coloured in red and inversely compounds with significantly lower levels in blue. It can be observed that in both ESI+ and ESI-differences in fatal outcome are more often based on increased levels.

Significance filtering based on volcano plot analysis reduced the number of metabolic features to 1987/ 935 (ESI+/- respectively) for severity and/or outcome combined. For those features, signal curation based on chromatogram and spectral quality (see Methods section), was performed in Compound Discoverer and a total of (526/409 ESI+/- respectively) features were retained. Table 2 below provides a breakdown of those compounds in terms of Metabolomics Standards Initiative (MSI) levels (Sumner et al., 2007), ranging from MSI level 1 to 4.

**Table 2.** Number of metabolic features remaining after LC-MS/MS data pre-processing, statistical selection of significant features and manual curation for chromatographic peak shape and spectral signal. Compounds where no molecular formula could be assigned by CD3.1 (see Methods) were discarded.

| MSI level (Sumner et al., 2007) | ESI+ | ESI- |
| --- | --- | --- |
| **Level 4: Molecular formula match** | 526 | 409 |
| **Level 3: Molecular formula + Mass library match** | 384 | 283 |
| **Level 2: all above plus MS/MS match (spectral match $\geq 70\%$)** | 45 | 33 |
| **Level 1: MS/MS experimental in-house library match** | 14 | 8 |

Evaluation of the four predictive models on this reduced set of 935 ESI+/- metabolic features showed the best generalization results using Bayesian logistics regression; therefore, all the following results are based on this model. The mean area under the curve (AUC) was calculated as 0.836 (SD 0.069) for severity and 0.807 (SD 0.081) for outcome, demonstrating good predictive power of the patient metabolome. However, a mass spectrometry-derived model with some 900 compounds is not a practical solution (Kenny et al., 2010) for an assay of general utility, and thus subsets of these compounds were investigated further. The sub-group selection was guided by metabolic pathway enrichment analysis and manual curation of well identified (i.e., MSI level 1 or 2) compounds with known biological relevance. A final model with 20 compounds selected in this way showed a reasonable cross-validated mean AUC of 0.793 (SD 0.080) for severity and 0.792 (0.090) for outcome. Mean balanced accuracy was calculated at 0.716 (0.088) and 0.655 (0.098) for severity and outcome, respectively. Representative receiver operating characteristic (ROC) curves for one random train-test split are shown in Figure 3.

**Figure 3.**
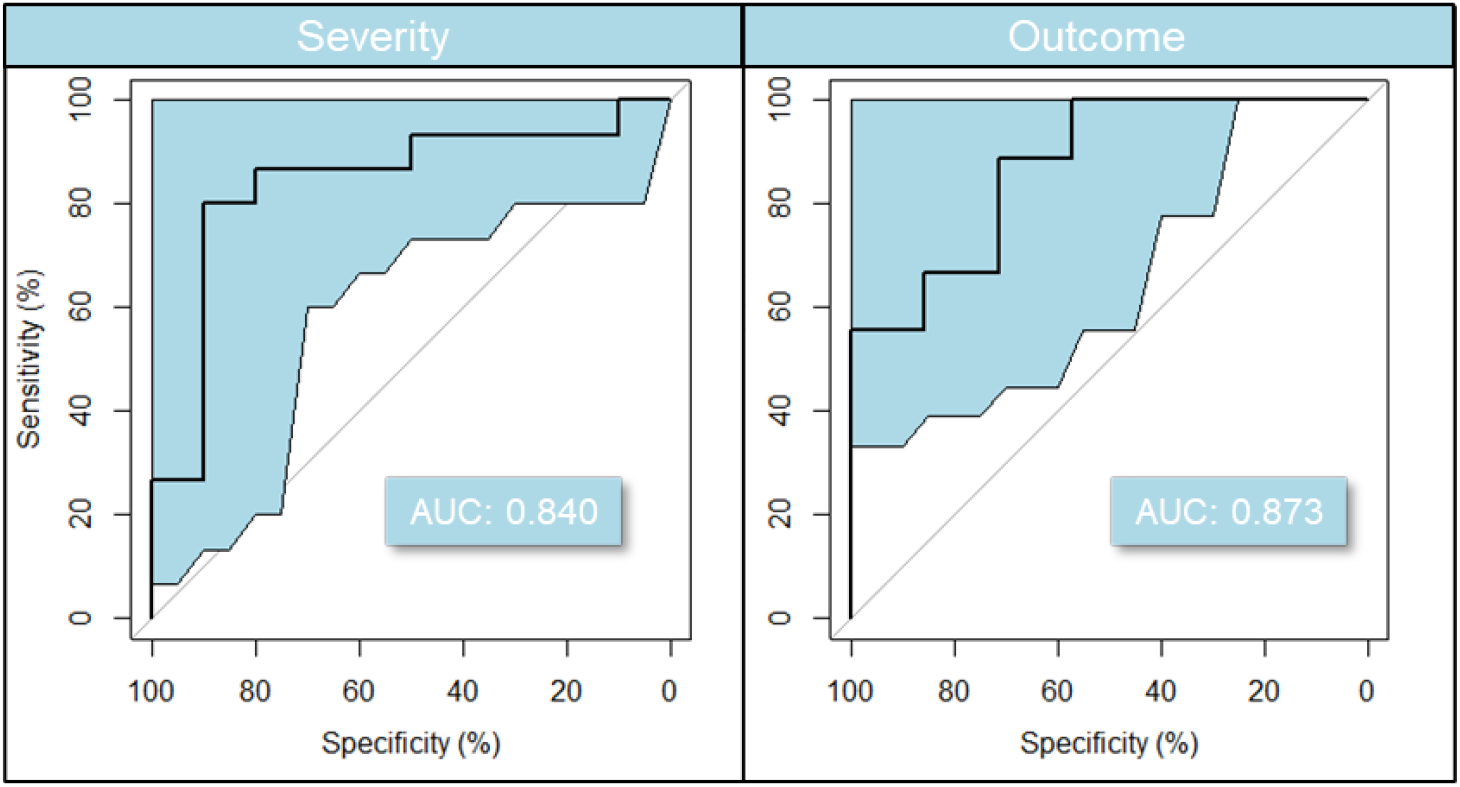
Predictive model based on 20 compounds selected for their identification confidence and known biological role (see Figure 4). Balanced accuracy and AUC for the represented in the graphic model are in the 0.70s and 0.80s, respectively. ROC 95% confidence intervals were calculated with 2000 stratified bootstrap replicates on the test data and are presented as blue shading around the mean curve. A Monte Carlo cross-validation results estimate the model balanced accuracy at 0.716 for severity and 0.655 for outcome. Cross-validated AUC was calculated at ∼0.79 in both conditions.

### Metabolic pathways linked to severity and outcome

Pathway enrichment analysis was performed with MUMMICHOG (Li et al., 2013) as implemented in MetaboAnalyst (Pang et al., 2020) as a way of selecting biologically relevant sub-groups of compounds. As expected, no ‘entire’ pathways showed significant *p*-values when ESI+ and ESI-results were grouped together (Anderson et al., 2014; Kell and Goodacre, 2014; Kell and Westerhoff, 1986). However, pathways that showed multiple significant hits were further investigated manually. These included pyrimidine metabolism and tryptophan metabolism. A number of these are in accordance with recently published studies comparing COVID-19 patients <u>to healthy controls</u> (Blasco et al., 2020; Overmyer et al., 2020; Thomas et al., 2020). The 20 compounds selected with this approach (shown with their significance in patient outcome Figure 4) are further discussed below. Selected compound significance in the prediction of COVID-19 severity is shown in Figure S4.

**Figure 4.**
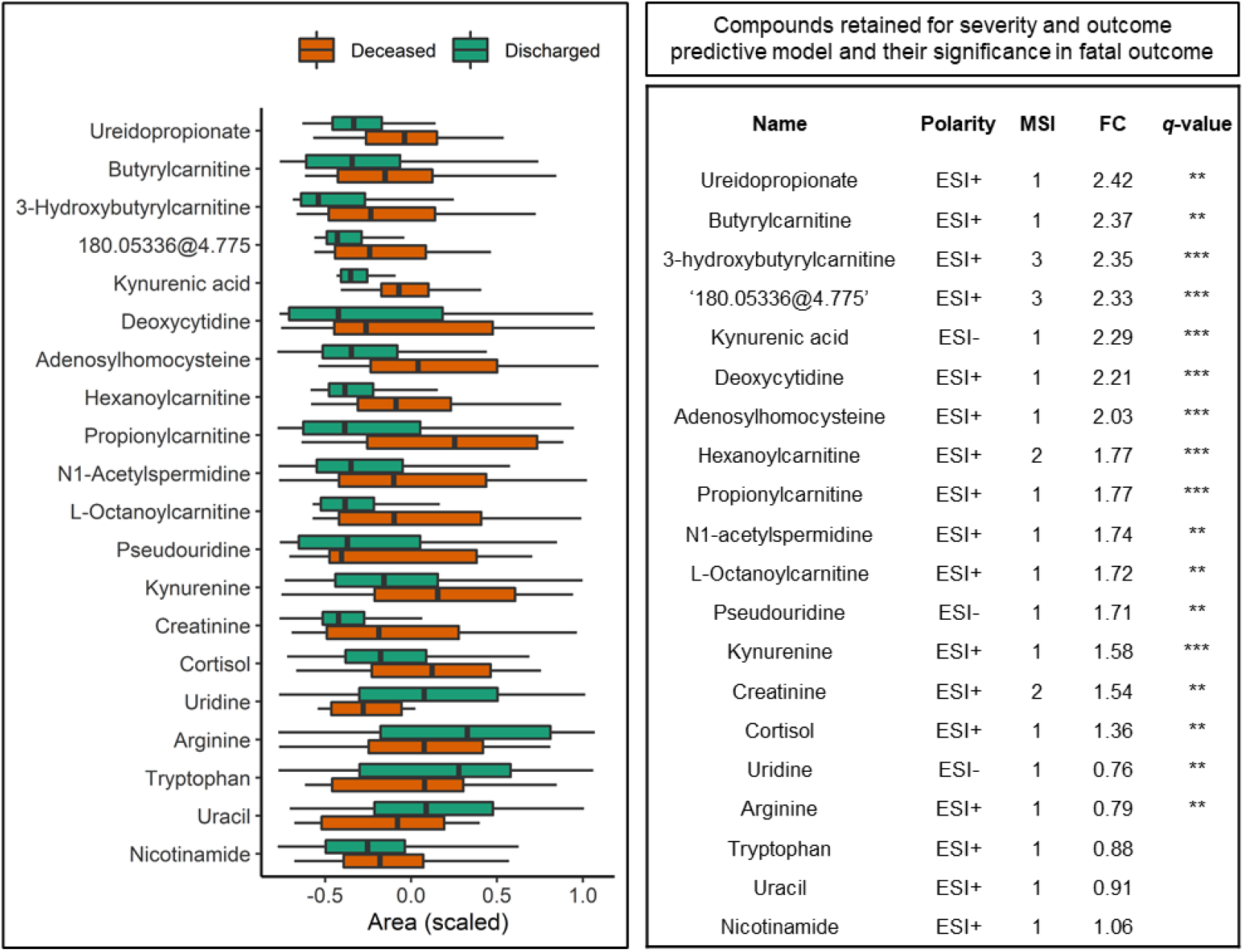
Compounds retained for severity and outcome predictive model. Box plot shows compound area differences between discharged and deceased patients ordered by fold change. Compound areas are standardized (mean = 0, SD = 1) to facilitate comparison. Boxes represent the quartiles Q1 to Q3 with Q2 (i.e., median) line in the middle. The ‘whiskers’ depict the upper and lower limit i.e., Q1 ± (Q3-Q1). For visualization simplicity the data is clipped between 10 and 90 percentiles. The table on the right side of the figure shows detailed information about the compounds including FC and q-value (false discovery rate corrected p-value) following ‘star’ notation i.e., ‘^***^’ correspond to q-values <0.001, ‘^**^’ <0.01, and missing when >0.1.

#### Elevated serum deoxycytidine and ureidopropionate association with severity and outcome

Deoxycytidine levels were increased over 2-fold in patient samples at admission who went on to develop severe symptoms or subsequently died (Figure 5 A and B). Furthermore, ureidopropionate, a pyrimidine degradation product, showed the strongest FC increase of 2.4. Similarly, elevated cytosine levels were found in COVID-19 patients measured against healthy controls and 15 days post infection in (Blasco et al., 2020), but severity or outcome were not discriminated as they were here.

**Figure 5.**
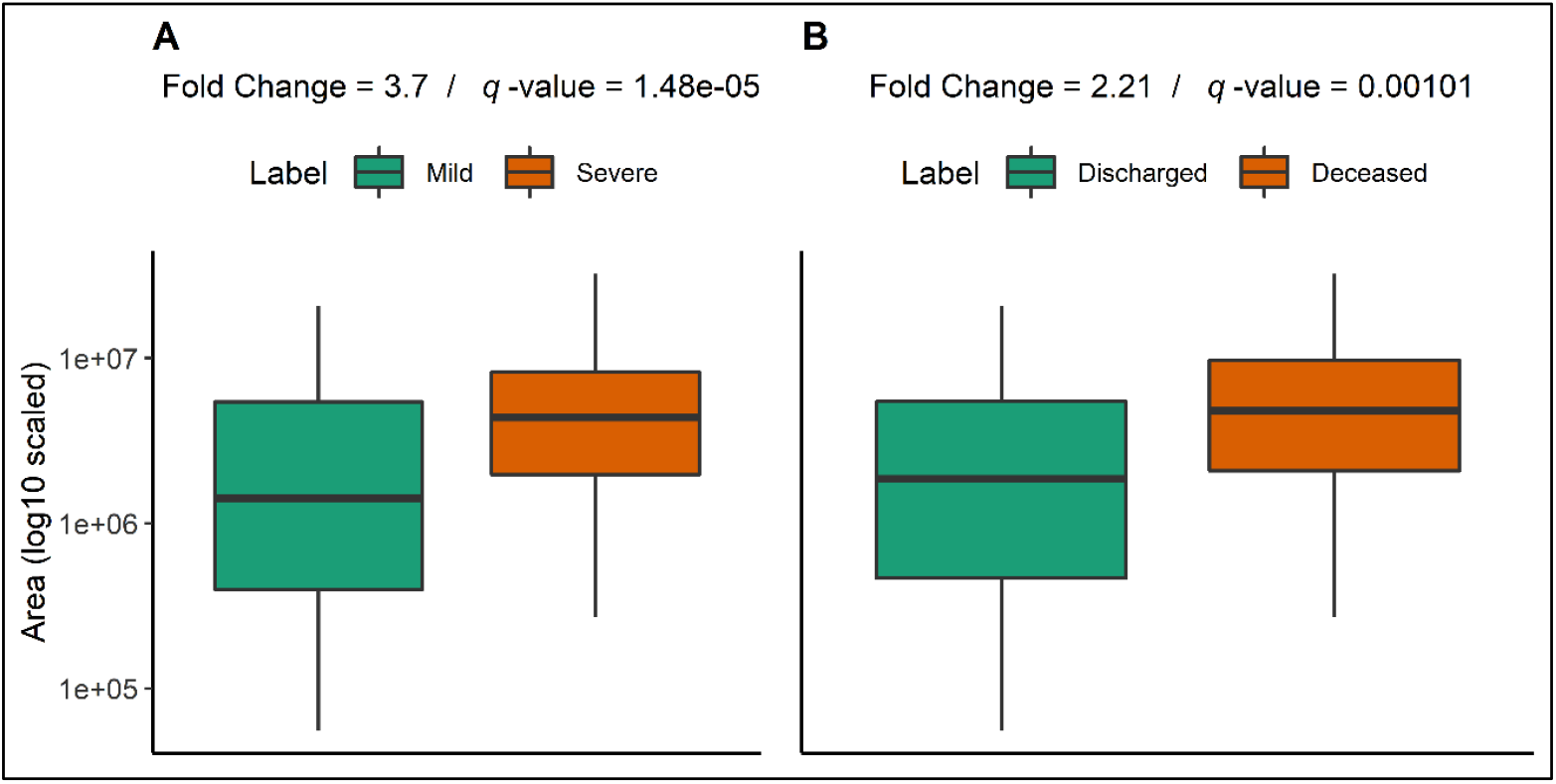
Deoxycytidine levels in patients according to (A) severity or (B) outcome. Boxplot markings follow the same standard as Figure 4.

Uridine, another pyrimidine, was found to be significantly decreased in fatal outcome cases; however, its FC was close, but not significant in severe cases. Moreover, pseudouridine, an isomer of uridine, was increased in patient samples with severe disease progression or deceased outcome. Pseudouridine (Figure 4) is a marker of cell ribosomal RNA (rRNA) turnover (Nakano et al., 1993), for instance in heart failure (Dunn et al., 2007).

#### Tryptophan and kynurenine metabolism compounds associate with both severity and outcome

Kynurenine (a tryptophan degradation product) was significantly increased in both severe cases and in patients who died, with a 1.5-fold change. The difference was even more marked for kynurenic acid in outcome, with levels increasing over 2-fold.

In addition to changes in kynurenine and kynurenic acid, our results showed that a reduction in levels of serotonin (MSI level 3) and melatonin (MSI level 3) were associated with COVID-19 severity and death. We also observed an increase in serum levels of cortisol with *q*-value of 0.006 and ∼1.3-fold in severe cases as well as those who died. The observed changes in tryptophan metabolism appear to be related to an immune response to SARS-CoV-2 since the activity of tryptophan dioxygenase, a tryptophan-degrading enzyme, is upregulated by cortisol and initiates degradation of tryptophan to kynurenine. Previous research (Thomas et al., 2020) has found a correlation between increases of interleukin 6 and kynurenine levels in COVID-19 patients compared to controls. Finally, kynurenine pathway upregulation as part of an inflammatory response is known to negatively impact serotonin levels (Hunt et al., 2020; Li et al., 2017), as was observed in our results.

Interestingly, we observed nicotinamide (MSI 1) levels to be 1.5-fold higher in severe cases, but not significantly changed with respect to outcome. It is important to note that nicotinamide is related to kynurenine/tryptophan metabolism (Murray, 2003).

#### Beta oxidation metabolites, acylcarnitines, increased in severe and deceased outcome patients

Multiple fatty acyl carnitines, e.g. hydroxybutyrylcarnitine (>2-fold), hexenoylcarnitine (1.4-fold) and hydroxyoctanoyl (∼2-fold), were found to be significantly higher in both severe and poor outcome cases. Long chain fatty acyl carnitines were not reliably detected here as the LC gradient used for data acquisition is optimized for more hydrophilic molecules. Therefore, fatty acyl carnitines with more than 10-carbon FA chains were excluded from the analysis. On the other hand, carnitine (MSI level 1) and its precursors, i.e., lysine (MSI 1) and methionine (MSI 1), were unchanged in both severity and outcome.

#### Other compounds

The final selection of compounds also includes arginine (MSI level 1) and *S*-adenosylhomocysteine (MSI level 2), a precursor to homocysteine. Levels of these compounds were significantly increased in patients who developed severe COVID-19 and those with a fatal outcome. Moreover, the model includes *N*1-acetylspermidine (MSI level 1) that was reported along with spermidine by (Thomas et al., 2020) as increased in COVID-19 patient sera compared to controls. Interestingly, in contrast to *N*1-acetylspermidine, spermidine did not show significant changes in relation to either severity or outcome. *N*1-acetylspermidine was also found related to COVID-19 prognostic in a smaller, recent study (Danlos et al., 2021); however, in contrast to our findings the lower levels were associated with an unfavourable outcome. Finally, the model includes creatinine (MSI level 2) which, in accordance with the clinically acquired data, tends to increase in patients with fatal outcomes but did not increase with disease severity.

### Model compounds adjusted for demographic factors and underlying conditions

Confounders are an important issue in omics studies (Broadhurst and Kell, 2006), and several factors (age, gender, BMI, and existing inflammatory diseases) are recognised as predisposing patients infected with COVID-19 to severe disease and poor outcome. Any impact on compound levels simply from population demographics and underlying conditions was explored with univariate logistic regression odds ratios (OR) and 95% confidence interval (95% CI) analysis. Individual compounds’ OR and 95% CI for outcome as adjusted for age, gender, BMI, diabetes, liver, kidney and cardiac disease and all together are presented in Table 3. The severity-based table is available in the supplementary information (Table S1). It is important to note that identifying links between those factors and the compounds of interest is meaningful when evaluating their biological role; however, being ‘confounded’ does not invalidate their predictive power and relevance in a predictive model. Additionally, it can be observed that not all compounds (e.g. kynurenic acid, cortisol) selected for their significant *q*-value and fold change showed significance based on 95% CI.

**Table 3.** Adjusted logistic regression results by outcome. Positive OR indicate increased levels in patients with poor outcome and are presented with OR (95% CI). Significance is presented in ‘star’ notation i.e., ‘ ^***^’ correspond to p-values <0.001, ‘ ^**^’ <0.01, ‘ ^*^’ <0.05, ‘.’ < 0.1 and missing when >0.1. Compounds are adjusted for age, gender, BMI, liver conditions, cardiovascular diseases, hypertension, kidney disease, diabetes and all together. Details of specific diseases in each category are available in the Methods section.

|  | Adjusted by: | None |  | Age |  | Gender |  | BMI |  | Liver disease |  | CV disease |  | Hypertension |  | Kidney disease |  | Diabetes |  | All |  |
| --- | --- | --- | --- | --- | --- | --- | --- | --- | --- | --- | --- | --- | --- | --- | --- | --- | --- | --- | --- | --- | --- |
|  | Compound | OR | Sig. | OR | Sig. | OR | Sig. | OR | Sig. | OR | Sig. | OR | Sig. | OR | Sig. | OR | Sig. | OR | Sig. | OR | Sig. |
| Nucleic acids metabolism | Deoxycytidine | 2.1<br>(1.4-3.6) | ** | 2<br>(1.2-3.4) | * | 2.3<br>(1.4-3.9) | ** | 2.1<br>(1.3-3.8) | ** | 2.1<br>(1.4-3.6) | ** | 2.1<br>(1.3-3.5) | ** | 2<br>(1.3-3.4) | ** | 2<br>(1.3-3.5) | ** | 2<br>(1.3-3.5) | ** | 2.5<br>(1.4-4.9) | ** |
|  | Uracil | 0.75<br>(0.47-1.1) |  | 0.84<br>(0.56-1.2) |  | 0.79<br>(0.49-1.2) |  | 0.83<br>(0.56-1.2) |  | 0.75<br>(0.47-1.2) |  | 0.72<br>(0.44-1.1) |  | 0.78<br>(0.48-1.2) |  | 0.79<br>(0.49-1.2) |  | 0.79<br>(0.49-1.2) |  | 0.81<br>(0.45-1.4) |  |
|  | Ureidopropionate | 2.8<br>(1.4-6.9) | * | 7.5<br>(2.8-24) | *** | 2.7<br>(1.4-6.5) | * | 3.2<br>(1.4-8.4) | ** | 2.8<br>(1.4-6.9) | * | 3<br>(1.5-7.3) | ** | 3.9<br>(1.8-10) | ** | 3.2<br>(1.5-8.3) | ** | 3.2<br>(1.5-8.3) | ** | 4.5<br>(1.8-14) | ** |
|  | Pseudouridine | 1.9<br>(1.3-2.8) | ** | 1.5<br>(0.97-2.4) | . | 1.9<br>(1.2-2.9) | ** | 1.6<br>(1.1-2.5) | * | 1.9<br>(1.3-2.8) | ** | 1.8<br>(1.2-2.8) | ** | 1.7<br>(1.1-2.7) | * | 2<br>(1.2-3.5) | * | 2<br>(1.2-3.5) | * | 1.6<br>(0.91-2.9) |  |
|  | Uridine | 0.39<br>(0.21-0.67) | ** | 0.58<br>(0.37-0.88) | * | 0.4<br>(0.22-0.7) | ** | 0.61<br>(0.39-0.91) | * | 0.39<br>(0.21-0.67) | ** | 0.36<br>(0.19-0.63) | *** | 0.41<br>(0.22-0.7) | ** | 0.41<br>(0.22-0.71) | ** | 0.41<br>(0.22-0.71) | ** | 0.41<br>(0.19-0.82) | * |
| Kynurenine metabolism | Tryptophan | 0.71<br>(0.46-1.1) |  | 0.88<br>(0.59-1.3) |  | 0.68<br>(0.43-1.1) | . | 0.79<br>(0.53-1.2) |  | 0.72<br>(0.46-1.1) |  | 0.72<br>(0.46-1.1) |  | 0.73<br>(0.47-1.1) |  | 0.76<br>(0.48-1.2) |  | 0.76<br>(0.48-1.2) |  | 0.72<br>(0.42-1.2) |  |
|  | Kynurenine | 2.7<br>(1.7-4.8) | *** | 2.3<br>(1.4-4.2) | ** | 2.8<br>(1.8-5.1) | *** | 2.3<br>(1.4-3.9) | ** | 2.7<br>(1.7-4.8) | *** | 2.9<br>(1.7-5.4) | *** | 2.6<br>(1.6-4.6) | *** | 2.6<br>(1.6-4.7) | *** | 2.6<br>(1.6-4.7) | *** | 2.6<br>(1.5-5) | ** |
|  | Kynurenic acid | 1.5<br>(0.99-2.6) |  | 1.4<br>(0.91-2.7) |  | 1.5<br>(0.96-2.6) |  | 1.4<br>(0.88-2.6) |  | 1.5<br>(0.98-2.6) |  | 1.5<br>(1-2.6) | . | 1.4<br>(0.91-2.4) |  | 1.3<br>(0.87-2.4) |  | 1.3<br>(0.87-2.4) |  | 1.2<br>(0.73-2.1) |  |
|  | Cortisol | 1.4<br>(0.93-2.2) |  | 1.3<br>(0.9-2.1) |  | 1.4<br>(0.93-2.2) |  | 1.2<br>(0.82-2) |  | 1.4<br>(0.94-2.2) |  | 1.4<br>(0.91-2.1) |  | 1.4<br>(0.92-2.2) |  | 1.4<br>(0.93-2.1) |  | 1.4<br>(0.93-2.1) |  | 1.4<br>(0.86-2.3) |  |
|  | Nicotinamide | 1.1<br>(0.68-1.5) |  | 1.2<br>(0.8-1.8) |  | 1<br>(0.67-1.5) |  | 1<br>(0.69-1.5) |  | 1<br>(0.67-1.5) |  | 1.1<br>(0.68-1.5) |  | 1.1<br>(0.69-1.6) |  | 1.1<br>(0.67-1.5) |  | 1.1<br>(0.67-1.5) |  | 1.2<br>(0.7-1.9) |  |
| Fatty acyl carnitines | Propionylcarnitine | 2.2<br>(1.5-3.6) | *** | 2<br>(1.3-3.2) | ** | 2.2<br>(1.4-3.5) | *** | 1.9<br>(1.2-3) | ** | 2.3<br>(1.5-3.6) | *** | 2.2<br>(1.4-3.5) | *** | 2.1<br>(1.4-3.5) | *** | 2.2<br>(1.4-3.5) | ** | 2.2<br>(1.4-3.5) | ** | 1.9<br>(1.1-3.2) | * |
|  | Butyrylcarnitine | 2.5<br>(1.6-4.4) | *** | 2.8<br>(1.6-5.7) | ** | 2.5<br>(1.6-4.5) | *** | 2.3<br>(1.3-4.4) | ** | 2.6<br>(1.6-4.6) | *** | 2.5<br>(1.5-4.4) | *** | 2.4<br>(1.5-4.3) | *** | 2.4<br>(1.5-4.3) | *** | 2.4<br>(1.5-4.3) | *** | 2.2<br>(1.3-4.4) | * |
|  | 3-hydroxybutyrylcarnitine | 1.9<br>(1.3-2.9) | ** | 1.7<br>(1.1-2.8) | * | 2.1<br>(1.4-3.3) | *** | 1.5<br>(0.98-2.4) | . | 1.9<br>(1.3-2.9) | ** | 1.8<br>(1.2-2.9) | ** | 1.8<br>(1.2-2.9) | ** | 1.8<br>(1.2-2.8) | ** | 1.8<br>(1.2-2.8) | ** | 1.6<br>(1-2.7) | * |
|  | Hexanoylcarnitine | 1.4<br>(0.95-2.4) |  | 1.2<br>(0.82-2.2) |  | 1.4<br>(0.92-2.3) |  | 1.3<br>(0.83-2.2) |  | 1.4<br>(0.95-2.4) |  | 1.4<br>(0.93-2.3) |  | 1.3<br>(0.9-2.3) |  | 1.3<br>(0.88-2.2) |  | 1.3<br>(0.88-2.2) |  | 1.1<br>(0.67-1.7) |  |
|  | L-Octanoylcarnitine | 1.4<br>(0.93-2.2) |  | 1.2<br>(0.78-1.9) |  | 1.3<br>(0.9-2.2) |  | 1.2<br>(0.79-1.9) |  | 1.4<br>(0.93-2.2) |  | 1.3<br>(0.91-2.2) |  | 1.3<br>(0.87-2.1) |  | 1.3<br>(0.84-2.1) |  | 1.3<br>(0.84-2.1) |  | 0.95<br>(0.56-1.5) |  |
| Other | Creatinine | 1.5<br>(1-2.4) | * | 1.4<br>(0.91-2.2) |  | 1.5<br>(1-2.3) | . | 1.4<br>(0.93-2.4) |  | 1.5<br>(1-2.4) | * | 1.6<br>(1.1-2.4) | * | 1.4<br>(0.93-2.2) |  | 1.4<br>(0.91-2.3) |  | 1.4<br>(0.91-2.3) |  | 1.4<br>(0.82-2.4) |  |
|  | Arginine | 0.53<br>(0.33-0.81) | ** | 0.66<br>(0.43-0.97) | * | 0.52<br>(0.32-0.81) | ** | 0.69<br>(0.46-1) | . | 0.51<br>(0.32-0.8) | ** | 0.53<br>(0.33-0.81) | ** | 0.53<br>(0.33-0.82) | ** | 0.55<br>(0.34-0.86) | * | 0.55<br>(0.34-0.86) | * | 0.59<br>(0.33-1) | . |
|  | N1-acetylspermidine | 1.9<br>(1.3-3) | ** | 1.8<br>(1.2-3.1) | * | 1.9<br>(1.3-3.1) | ** | 1.7<br>(1.1-2.9) | * | 2<br>(1.3-3.2) | ** | 1.9<br>(1.2-3) | ** | 1.9<br>(1.3-3.1) | ** | 1.8<br>(1.2-2.9) | ** | 1.8<br>(1.2-2.9) | ** | 1.7<br>(1-3) | * |
|  | '180.05336@4.775' | 1.6<br>(1.1-2.6) | * | 1.5<br>(1-2.8) | . | 1.6<br>(1.1-2.5) | * | 1.6<br>(1-2.9) | . | 1.7<br>(1.1-2.7) | * | 1.6<br>(1.1-2.6) | * | 1.5<br>(1-2.4) | * | 1.5<br>(1-2.6) | . | 1.5<br>(1-2.6) | . | 1.5<br>(0.92-2.6) |  |
|  | Adenosylhomocysteine | 2.3<br>(1.4-4.2) | ** | 2.2<br>(1.3-4.5) | * | 2.3<br>(1.4-4.4) | ** | 2.3<br>(1.3-4.7) | * | 2.3<br>(1.4-4.2) | ** | 2.2<br>(1.4-4.1) | ** | 2.2<br>(1.3-4.1) | ** | 2.3<br>(1.3-4.8) | * | 2.3<br>(1.3-4.8) | * | 2.1<br>(1.2-4.3) | * |

An important result from this analysis is that cytosine and kynurenine remained significant regardless of patient demographics or underlying conditions. By contrast, increases in abundance of short and medium fatty acylcarnitines in severity and outcome appear to be partially explained by age and BMI, as OR tend to decrease when adjusted; this may be correlated to frailty in ageing (Rattray et al., 2019). After adjusting for all recorded conditions in the study, only butyrylcarnitine, 3-hydroxybutyrylcarnitine and hydroxyhexanoylcarnitine showed a strong relation to disease severity. In respect to outcome, butyrylcarnitine remained the most significant. This indicates that higher levels of acyl carnitines are likely linked to metabolic differences in the patients prior to the viral infection. As such they could potentially indicate a risk group. Moreover, pseudouridine’s significance was impacted by age, BMI and mildly by cardiac conditions and hypertension. This in itself is not a confounding issue since both age and BMI also contribute statistically to the outcome of COVID-19 (Hussain et al., 2020; Iaccarino et al., 2020).

Interestingly ureidopropionate, despite maintaining strong significance when adjusted for all factors, showed an increased OR when adjusted for age in fatal outcome case. In contrast, ureidopropionate’s OR dropped when adjusted for age and BMI in severe (vs mild) cases. Lower ureidopropionate levels have also been associated with an increased risk of developing type 2 diabetes mellitus and coronary artery disease (Ottosson et al., 2018).

A recent publication found an association between gender and kynurenic acid levels in COVID-19 patients suggestive of sex-specific differences in immune responses and clinical outcomes (Cai et al., 2021). However, in our results gender correction did not impact kynurenic acid OR in either outcome or severity.

### Validation study

Validation is an important aspect in metabolomics studies and as well as this being statistical, more importantly is replication of the results (Gromski et al., 2015). A Bayesian logistic model was trained on the discovery group of patients and used to predict the validation group of separate patients in a blind manner i.e., data acquisition and data analysis (model predictions) were performed without labels knowledge. The prediction results showed in Figure 6 evaluated a ROC AUC of 0.83 (CI 0.74 – 0.91) for outcome and 0.76 (CI 0.67 – 0.86) for severity.

**Figure 6.**
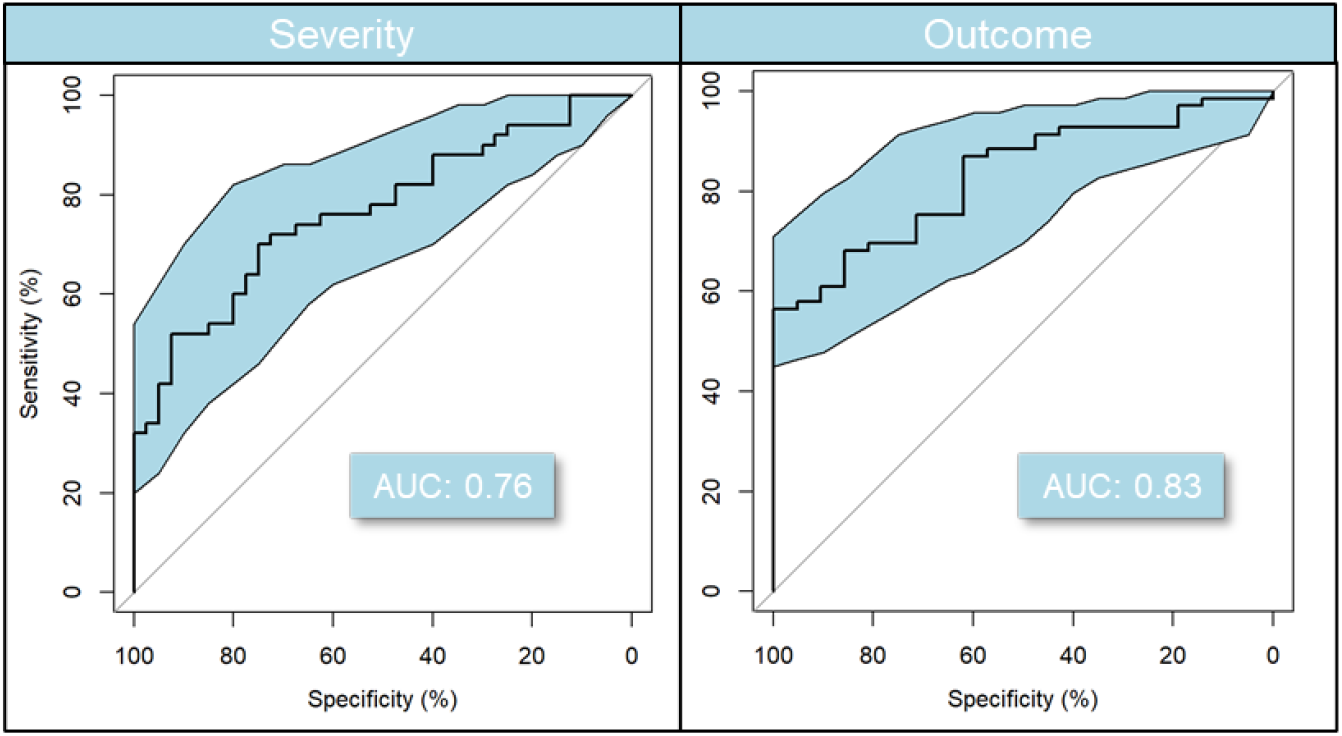
Predictive model performance based on 17 ESI+ compounds previously selected in the discovery study. ROC 95% confidence intervals were calculated with 2000 stratified bootstrap replicates on the test data and are presented as blue shading around the mean curve. A Monte Carlo cross-validation results estimate the model balanced accuracy at 0.63 for severity and outcome.

The validation study effort was focused on the 20 compounds identified in the discovery study. LC-MS/MS peak areas were extracted for these compounds in Trace Finder (see Materials and Methods) to allow the best results in signal mapping between batches and studies. In the case of ‘deoxycytidine’ all three cytosine events were summed up i.e., cytosine, cytidine, deoxycytidine. This approach was employed because automatic software pre-processing in Compound Discoverer showed multiple alignment issues between batches of those cytosine events close in RT and resulting from cytosine, cytidine, and deoxycytidine source fragmentation (see Sup Section 1.1). For this reason, these will be referred to as cytosine-based nucleosides in the following paragraphs.

Kynurenic acid (ESI-) showed poor acquisition signal in the validation study and was removed from the model. Uridine (ESI-) and pseudouridine (ESI-) showed significant distribution differences after normalization between the data acquired for the discovery study patients and validation study patients. Such study bias can mislead the model therefore these two compounds were also removed from the validation model.

All areas for the remaining 17 ESI+ compounds were normalized by batch and between batches for all discovery and validation batches. This ensured relative areas of the 17 compounds were comparable between the 2 studies so the model could be trained on the discovery study and used to predict the validation study patients.

It is important to note that at the time of the validation cohort, which was later than those in the discovery phase, patients were regularly treated with dexamethasone, an anti-inflammatory corticosteroid,(NHS, 2020; RECOVERY, 2021) that by this time was known to have efficacy as a treatment agent. This change in patient treatment is likely a possible explanation of the drop in AUC for severity prediction as this could have influenced the patient state severity perception.

As shown in Figure 7, higher levels of *S*-adenosylhomocysteine, propionylcarnitine, ureidopropionate and kynurenine are highly influential factors in this multivariate outcome model, combined with low levels of tryptophan and uracil. Interestingly creatinine, octanoylcarnitine and hexanoylcarnitine show a negative influence once conditioned on these other compounds. In terms of severity, ureidopropionate and *S*-adenosylhomocysteine showed the strongest positive influence. However, these results need to be interpreted with caution due to conditioning e.g., on mediators. Still, it is interesting to note that there is broad agreement on directionality between the models.

**Figure 7.**
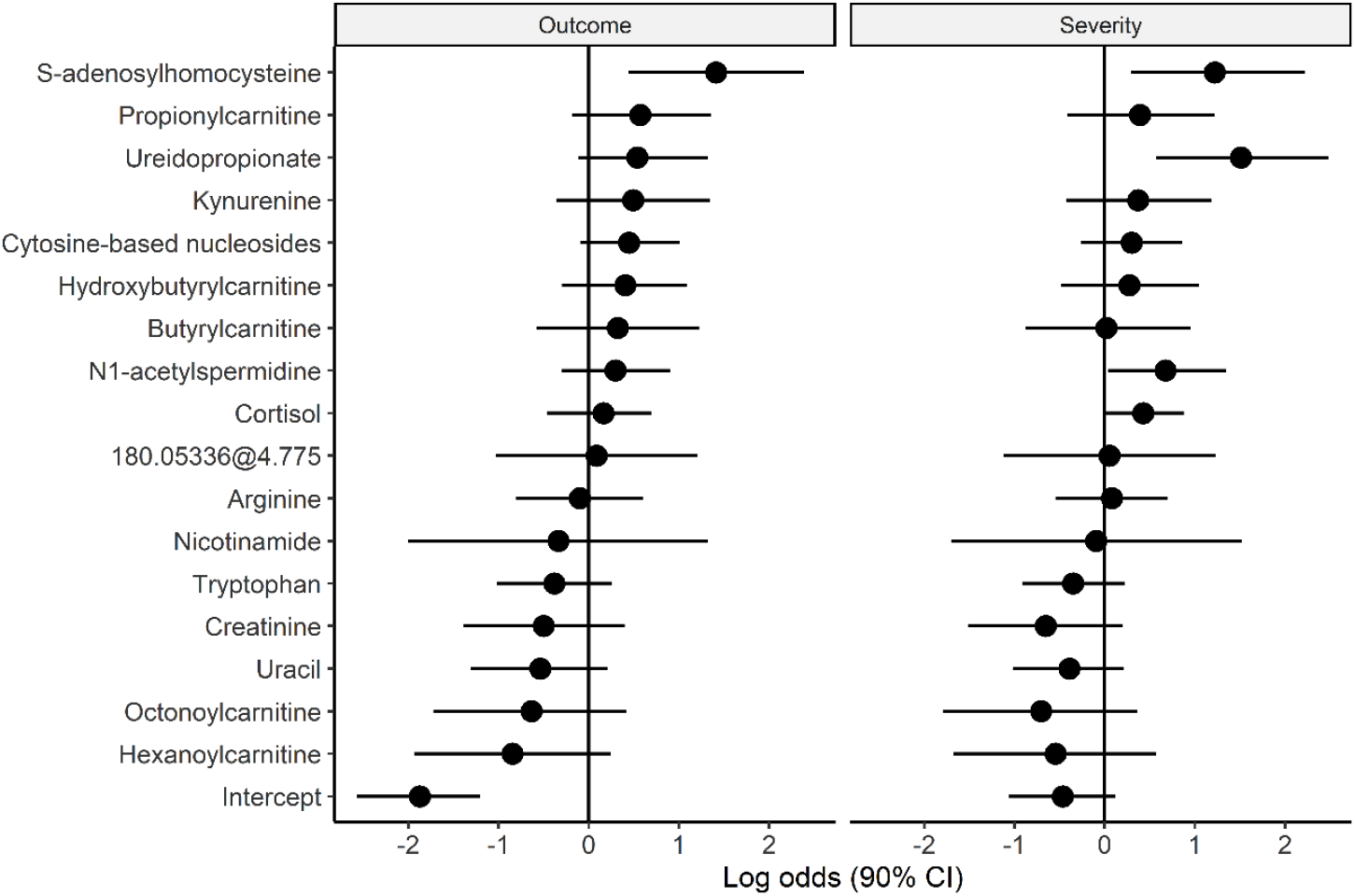
Plot of log odds with 90% posterior CI for both outcome and severity model ordered by mean log odds in outcome. These parameters are from the multivariate model and the log odds CI are not meant to indicate individual significance, but to illustrate alignment in directionality between outcome and severity models. Intercept in the figure refers to the intercept of the generalized linear model, in this case the log odds when all predictors are equal to their mean as data is standardized (mean = 0, SD = 1).

### Longitudinal study

Finally, a longitudinal sample of 28 severe patients with divergent outcomes from the discovery cohort (13 deceased and 15 discharged) was also analysed using LC-MS in 7 batches for a total of 198 samples. The samples were acquired through the patients’ stay on different days and with a different number of samples for each patient. Areas of the compounds previously selected in the predictive model were extracted in TraceFinder to allow for reliable comparison as alignment between batches suffered similar issues as those described in the validation study section.

Some preliminary results of the longitudinal data in form of a line plot of the time evolution of the measured compounds did not seems to display a clear patten. This is most likely due to the irregular interval between samples, the different times of day of sample acquisition, medication interference, measurements variability and the general complexity of biological processes. However, when focusing the analysis on the changes of those compounds throughout the hospital stay, i.e., the difference between the first and last sample of each patient, some compounds showed significant directional changes via logistic regression analysis. Figure 8 shows the significant compounds, ureidopropionate (A), uracil (B), arginine (C) and tryptophan(D).

**Figure 8.**
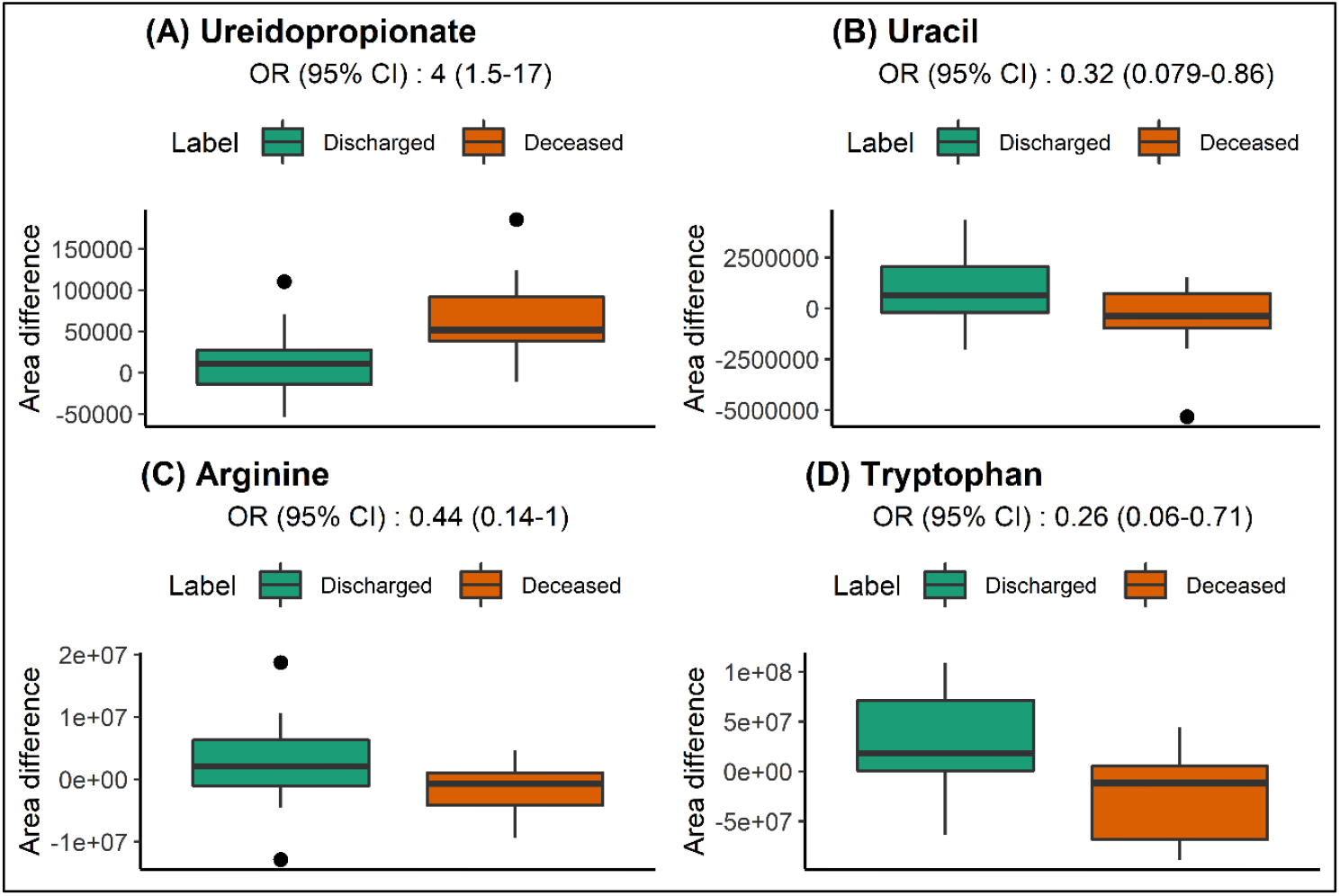
Level of selected compounds for significant change in level between first and last patient sample. Ureidopropionate (A), uracil (B), arginine (C) and tryptophan (D) levels showed significantly different evolution measured by logistic regression OR and 95%CI. Boxplot markings follow the same standard as Figure 4.

It appears that levels of ureidopropionate are significantly increased in deceased patients but they have stayed relatively similar in discharged patients. In addition, the role of this pathway is strengthened by the significant inverse relation in uracil levels (an increase) in discharged patients. Interestingly, cytosine-based nucleotides did not show significant differences in levels between deceased and discharged patients 0.67 (0.26-1.5) OR (95% CI), with the wide CI indicating strong variability between individuals. When it comes to tryptophan levels, we can see that discharged patients experienced increases in levels, where deceased patients saw a drop in tryptophan levels. Neither kynurenine 0.66 (0.27-1.4) or kynurenic acid 1.2 (0.52-3.6) showed significant deltas probably due to the broad variation between patients or possibly suffering from precursor deficiency in some cases.

## Discussion

In this study we employed untargeted metabolomics using LC-MS/MS to detect and measure changes in the baseline serum metabolome of a cohort of 120 COVID-19-infected patients at the point of hospital admission. We than validated our findings in a blind study on an additional 90 patients and explored the compounds’ temporal evolution in a longitudinal data of 28 severe patients. Our aim was to find not only prognostic markers of subsequent disease severity and outcome, but to also understand what effects COVID-19 infection has on the patient’s metabolome, and conversely what effect patient biochemistry and physiology might have on infection development. These results could then be used to guide patient treatment and medical attention requirements following COVID-19 diagnosis. Most early studies applying metabolomics to COVID-19 human patients have compared them against healthy controls; this is of limited predictive value, as rapid PCR and PoC antibody tests exist, and does not provide an insight into what biochemical changes drive disease severity or outcome. Moreover, a number of these studies (Table 1), used either a small patient cohort or applied targeted metabolomics restricting the breath of possible findings; some also lacked proper statistical analysis (see (Broadhurst and Kell, 2006; Trivedi et al., 2017)).

Our results showed that distinct alterations in the serum metabolome were already capable of identifying patients with higher risk of severe illness or fatal outcome at the time of diagnosis and admission. More importantly, using both univariate and a multiple predictor Bayesian logistic regression model we found a subset of 20 metabolites (16 of which could be identified to MSI Level 1) where most of them have relevant biological functions that are predictive of subsequent disease severity and patient outcome with AUC 0.792 and 0.793 respectively. These changes centred particularly around pyrimidine, tryptophan and acylcarnitine metabolism, which can be related to viral presence/replication (and/or the virus’s stimulation of host biosynthesis), host inflammation response and alterations in energy metabolism.

### Alterations in pyrimidine metabolites predictive of severity and outcome

Viral infections induce characteristic changes in host cell metabolism to enable effective viral replication (Thaker et al., 2019). Moreover, the resulting metabolic impact and cellular reprogramming varies between viruses (even within the same family) and host cell type. In the case of SARS-CoV-2, cytosine has been described as pivotal in the virus’ evolution (Danchin and Marlière, 2020) where avoidance of host defence mechanisms have favoured a reduced cytosine proportion in viral RNA, estimated at 17.6%. When compared to typical human RNA, the cytosine proportion is significantly lower. In contrast to cytosine, the proportion of uracil in SARS-CoV-2 is estimated at around 32.4% (Danchin and Marlière, 2020) and it is significantly higher than that in human RNA. This difference between host nucleotides anabolic processes and viral RNA needs could result in significant levels of cytosine-based ribonucleotides and deoxyribonucleotides being accumulated in infected cells.

CTP synthetase, the enzyme converting UTP to CTP, is allosterically inhibited by CTP, therefore allowing the virus to thrive despite its RNA composition differences. However, the reduction of the ribonucleotides to deoxyribonucleotides by ribonucleotide reductase is nonspecific, and its activity is not modulated by CDP or CTP (Hofer et al., 2012). This could allow the small differences in CTP to UTP ratios to be amplified in their reduced form, consistent with the principles of metabolic control analysis (Kell and Mendes, 2012). The accumulation of CDP and CTP in infected cells could result in breakdown and potentially deoxycytidine being excreted from the cells as waste product or released at cell death. It is important to note that SARS-CoV-2 has been found to use lysosomes trafficking to exit the cell (Ghosh et al., 2020), therefore leaving an open question on how the deoxycytidine is being released in the systemic circulation.

The high levels of ureidopropionate, a product of pyrimidine catabolism, observed in patients with severe disease and poor outcome also show activation of salvage pathways, most likely driven by an excess of nucleosides. It is important to note that the first step of cytidine breakdown is the conversion to uridine by cytidine aminohydrolase therefore, closing the gap between cytidine / uridine levels.

Cytosine has been reported previosuly to discriminate between COVID-19 infection compared to uninfected individuals (Blasco et al., 2020), but has not been assessed regarding severity or outcome. Here, our analyses showed that deoxycitidine levels were predictive of subsequent disease severity and outcome; serum deoxycitidine levels were increased over 2-fold in serum from patient samples at admission who went on to develop severe symptoms or subsequently died (Figure 5 A and B). Given the reduced incorpration of cytosine into SARS-CoV-2, these results may indicate a higher viral load and replication, subsequently leading to the development of severe symptoms, in some cases resulting in death.

Increased viral reproduction activity could be due to an initial higher viral load, a host environment favourable to viral reproduction, or the effective stage of the infection. It is thus not possible to draw mechanistic conclusions from our results; nevertheless, the deoxycitidine OR remained stable when adjusted for demographic factors and known underlying conditions indicting that those factors are not significant with respect to viral replication rates. This would be more consistent with the fact that the initial load before admission is the most important parameter affecting both disease severity and outcome.

Given these results, measurement of deoxycitidine and ureidopropionate, could potentially allow tracking of viral activity and predict recovery or aggravation. However, as highlighted by (Migaud et al., 2020) the levels of certain nucleobases are also increased in patients who die from sepsis and acute respiratory failure. Further investigation would be required to tease out the contributions of SARS-CoV-2 viral replication to the levels of these pyrimidines against the secondary effects of the virus on the host (human) health.

Pseudouridine, was also noted to be increased in severe cases and poor outcome. As mentioned previously, pseudouridine (Figure 4) is a marker of cell (rRNA) turnover (Nakano et al., 1993), for instance in heart failure (Dunn et al., 2007). When adjusting for cardiovascular disease this compound remained significant but showed slight correlation with hypertension and age. Finally, when adjusted for all factors pseudouridine 95% CI lost significance indicating correlation with more than one factor. Interestingly, growing evidence points toward complications in COVID-19 arising through a vasculopathy and coagulopathy elicited by the infection (Pretorius et al., 2020) and may be indicative of this process.

### Tryptophan - kynurenine degradation

The degradation of tryptophan to kynurenine is often associated with increased inflammatory processes (Schröcksnadel et al., 2006). In the context of this study the non-significant decrease of tryptophan in severe cases could be explained either by a change in dietary habits, possible weight loss, sarcopenia, or more likely by the higher stimulation of tryptophan to kynurenine degradation indicated by the significant increase in levels of kynurenine and kynurenic acid. Upregulation of this process was further confirmed by the higher levels of cortisol, which stimulates tryptophan degradation, especially in severe cases (Table 3). An increase in patient kynurenine levels has also been reported in COVID-19 compared to controls in (Thomas et al., 2020), associated with severity in (Overmyer et al., 2020), and also linked to fatal sepsis development in (Migaud et al., 2020). This provides strong evidence of higher levels of immune response in severe cases and those with a fatal outcome. More recently, upregulation of tryptophan metabolites including kynurenine has been found to play a protective role in radiation injury during cancer radiotherapy (Guo et al., 2020) indicating that these relationships are more complicated than previously thought, and also involve the gut microbiome.

Furthermore, changes in levels of nicotinic acid reflected by its condensation product nicotinuric acid could possibly reflect a dysfunctional energy metabolism. Interestingly our data indicates increased levels of nicotinamide in severity and nicotinuric acid in outcome. Increased levels of nicotinuric acid in urine have been previously reported as pathogenic markers in metabolic syndrome and cardiovascular disease (Huang et al., 2013), again, consistent with the known cardiovascular effects of SARS-CoV-2 infection (Fox et al., 2020; Grobler et al., 2020; Leisman et al., 2020; Libby and Lüscher, 2020; Paranjpe et al., 2020; Pretorius et al., 2020; Zheng et al., 2020).

### Beta oxidation

Interestingly, levels of short and medium chain acylcarnitines were previously reported by (Thomas et al., 2020) as reduced in COVID-19 patients *versus* controls irrespective of Interleukin 6 (IL6) levels. In this study we identified multiple short chain acyl carnitines as increased in severe cases and those with a fatal outcome compared to mild cases and discharged patients. Changes in serum acylcarnitines have been previously associated with cardiovascular disease, diabetes and inflammation (Anderson et al., 2014).

Moreover, increased levels of octanoyl-l-carnitine have been previously associate with arterial stiffness (Kim et al., 2015) and dysregulation of the carnitine shuttle. Long chain acyl carnitines, not detected in this study, have been additionally reported in relation to frailty in the elderly (Rattray et al., 2019).

Indeed, adjusted logistic regression results (Table 3) here show that some of their significance can be explained by BMI levels. This could possibly indicate different levels change in energy metabolism as response to viral infection linked to preexisting phenotype; i.e., BMI and therefore represent a high-risk group.

### Compounds requiring further investigation

Our study highlighted a number of other compounds (not discussed here) that changed significantly in severity and outcome. However, as is common in metabolomic studies (Blaženović et al., 2018; Salek et al., 2013; Shrivastava et al., 2021), these require further rigorous identification following Metabolomics Standards Initiative reporting for chemical analysis (Sumner et al., 2007) and so were not included in the predictive model at this stage. Examples of such compounds include pentahomomethionine (MSI 3) and trihomomethionine (MSI 3), both sulfur-containing amino acids. These were both increased in severe cases and were especially high in patients with a fatal outcome. Other sulfur-containing compounds such as cysteine and taurine have been found to be reduced in COVID-19 cases compared to controls, and in COVID-19 patients with moderate-high IL-6 levels (Thomas et al., 2020). Homomethionines such as penta- and tri-homomethionine identified in our results are formed by transamination of oxo-acids that are themselves formed in fatty acid breakdown, possibly indicating a pro-catabolic phenotype as is common in inflammation (Underwood et al., 2006) and possibly suggestive of sarcopenia.

Another compound worthy of further attention is ergothioneine. Ergothioneine is a potent exogenous antioxidant, usually acquired via the consumption of mushrooms (Borodina et al., 2019; Cheah and Halliwell, 2012). The potential protective value in SARS-CoV-2 infections of ergothioneine was recently reviewed by (Cheah and Halliwell, 2020). The results of our study found ergothioneine levels to be following the expected trend, i.e., lower in severe cases and poor outcome in accordance with finding by (Wu et al., 2020); however its *q*-values (in a population whose mushroom consumption was neither monitored nor controlled) fell just slightly short of significance (see Figure S5 A and B). Moreover, piperine (MSI 2) (representative of black pepper consumption) was interestingly found significantly decreased in severe cases and poor outcome (see Figure S5 C and D). This most likely reflects dietary changes in the patients experiencing severe symptoms; however, the potential impact of piperine to the host organism is not fully understood.

### Limitations and future work

Whilst untargeted LC-MS analysis allows detection of a large number of compounds of diverse chemical classes, limitations in the compound coverage can result from sample preparation methods, LC solvents and gradient. In this study the serum extraction and LC gradient were targeted at hydrophilic compounds, therefore numerous lipids were not reliably measured, i.e., long chain acyl carnitines. Despite this limitation several lipids that exhibit amphiphilic properties e.g., phospholipids and fatty acids were detected and showed significant changes between our patient groups. However, confirmation of their precise identity will require further work before being integrated into a predictive model.

Additional limitations in the presented work come from the preselection of metabolic features that were individually significant in volcano analysis. While this simple method of feature selection narrows the list of compounds it also prevents us from identifying more complex interactions in the case of disjoint populations. More importantly such variable selection performed on all data require validation in a separate patient cohort.

Despite these limitations, multiple significant biological processes were identified as key in discriminating between disease severity and outcome. Future work will aim to validate these findings against a new patient cohort.

## Conclusions

We have here performed a well-powered, untargeted metabolomics analysis of serum of COVID-19 patients with the aim of finding prognostic markers of disease severity and outcome. Using both univariate and a multiple predictor Bayesian logistic regression model we found a subset of 20 metabolites with relevant biological functions that are predictive of subsequent disease severity and patient outcome. Although, no individual metabolite appeared be *strongly* discriminative on its own, a combined model based on viral activity, host immune response and underlying metabolic differences showed promising predictive results with AUC 0.792 and 0.793, for outcome and severity, respectively. Furthermore, we validated those finding in a blind study on 90 additional patients with AUC of 0.83 and 0.76. Longitudinal exploration of the potential markers some showed strong relation to poor outcome opening opportunities to differentiate between recovery and aggravation states. These markers hold promise to improve patient care upon COVID-19 infection and diagnosis. Building on these encouraging results, further work will aim explore in depths the longitudinal aspect of the disease and potential treatment indications of those findings.

## Methods

All LC-MS data acquired is freely available in mzML format in the MetaboLights repository (Haug et al., 2019) with study identifier MTBLS2997 (www.ebi.ac.uk/metabolights/).

### Sample acquisition

All samples were acquired at the Royal Liverpool University Hospital (RLUH) on the first positive SARS-CoV-2 test (not fasted and different times of the day). Surplus serum was saved after routine diagnostic testing on patients admitted to the hospital who subsequently tested positive for SARS-CoV-2 via PCR. Blood was initially collected into VACUETTE Clot Activator tubes (Greiner, Germany) within approximately 48 h of presentation and centrifuged at 1,500 x *g* for 10 min within 60 min of collection. Surplus serum was stored at –80 °C prior to processing and analysis. Ethical approval for the use of serum samples and associated metadata in this study was obtained from North West – Haydock research ethics committee (REC ref: 20/NW/0332).

Severity scoring was based on the level of respiratory support required and overall patient outcome where severe corresponds to required fraction of inspired oxygen (FIO_2_) > 40% and/or required CPAP and/or required invasive ventilation and/or did not survive. Patients were also stratified using the 4C Mortality Score (Knight et al., 2020). Patient demographics by severity and outcome are presented in Table 4, where mild cases encompass mild to moderate disease severity.

**Table 4.** Patient demographics by severity and outcome. Cohort demographics are presented in counts with percentage (%) of the group total or mean with interquartile range (Q1 – Q3) depending on the data nature. *P*-values indicating significant differences between groups follow ‘star’ notation i.e., ‘^***^’ correspond to p-values <0.001, ‘ ^**^’ <0.01, ‘ ^*^’ <0.05, ‘.’ < 0.1 and missing when > 0.1. P-values were not calculated for the group counts (N). This study looked at the serum samples from 120 COVID-19 patients. 49 patients developed severe symptoms and 31 patients died as a result of the infection. Severity score metrics based on the 4C Mortality score (Knight et al., 2020) are provided as group means. Some disparity can be observed in gender as women represent 13% of the severe cases and only 29% of the deceased patients. Age groups of severe and deceased patients also tend to be slightly higher. BMI do not show significant difference between groups. O_2_ support indicates the number of patients that required oxygen support at any time during their hospitalization. Mechanical respiratory support indicates the need of invasive support or continuous positive airway pressure (CPAP). Max FiO_2_ represents the maximum fraction of inspired oxygen required by the patient during the hospitalization period, where respiratory support capture the patients requiring any support at diagnosis and FiO_2_ represents the fraction of inspired oxygen required at time of sample acquisition. As expected, oxygen need and inspired fraction, are highly correlated with severity and fatal outcome. National Early Warning Score (NEWS) also showed correlation with both severity and outcome. Cardiac disease refers to multiple cardiovascular conditions, most frequently: ischemic heart disease, atrial fibrillation, and heart failure. Kidney disease is a grouping of stages G2 to G5 of chronic kidney disease as defined by the National Institute for Health and Care Excellence (NICE) (NICE, 2015). Liver disease in most cases refers to cirrhosis and hepatitis. Malignancy cases vary from lung, bladder, prostate, skin cancer to haematological. Those underlying conditions did not show significance in severity or outcome. Despite kidney disease classification not showing correlation, estimated Glomerular Filtration Rate (eGFR) levels showed significance in severity and outcome. Fever (temperature ≥ 38 °C), cough and shortness of breath (SOB) were noted at time of sample acquisition and did not show relation to severity or outcome. Pulse, systolic blood pressure (BP) and respiratory rate also taken at samples acquisition showed correlation with severity and outcome, where higher pulse, higher respiratory rate and lower blood pressure are associated with severe cases and poor outcome. Haemoglobin levels (Hb), white blood cell count (WBC), lymphocyte count (Lymphs), platelets count (PLTs), haematocrit (HCT) and alanine aminotransferase (ALT) measured at sample acquisition did not show significant correlation with severity and outcome. Urea, creatinine, and C-reactive protein (CRP) concentrations are consistently elevated in severe and deceased patients. Hb, WBC, Lymphs, PLTs and HCT were measured on Beckman analyser. ALT, urea, creatinine and CRP were measured on a Roche analyser. Reference ranges are provided in [] when available.

| Condition | Severity |  |  | Outcome |  |  |
| --- | --- | --- | --- | --- | --- | --- |
|  | Mild | Severe | P-v | Discharged | Deceased | P-v |
| **N** | 71 | 49 | - | 89 | 31 | - |
| **4C score** | 8.39 ( 4.75 - 12) | 12.87 (10.00 - 16) | *** | 8.91 ( 5.25 - 12) | 14.34 (13.00 - 16) | *** |
| **Male** | 38 (54%) | 32 (65%) |  | 48 (54%) | 22 (71%) | . |
| **Female** | 33 (46%) | 17 (35%) |  | 41 (46%) | 9 (29%) | . |
| **Age** | 66.0 (54 - 77) | 69.2 (56 - 87) |  | 63.8 (52.0 - 77.0) | 77.1 (65.5 - 88.5) | *** |
| **BMI** | 26.4 (21.8 - 28.6) | 27.0 (23.0 - 32.0) |  | 26.7 (22.3 - 29.0) | 26.4 (21.9 - 29.6) |  |
| **O<sub>2</sub> support** | 32 ( 45%) | 49 (100%) | *** | 50 ( 56%) | 31 (100%) | *** |
| **Mechanical respiratory support** | 1 (1%) | 12 (25%) | *** | 3 (10%) | 10 (11%) |  |
| **Max FiO<sub>2</sub>** | 24.7 (21 - 28) | 71.6 (40 - 100) | *** | 33.3 (21 - 35) | 74.2 (36 - 100) | *** |
| **Required O<sub>2</sub> on presentation** | 17 (24%) | 39 (80%) | *** | 34 (38%) | 22 (71%) | ** |
| **FiO<sub>2</sub> (%) on presentation** | 22.8 (21 - 21) | 47.0 (28 - 60) | *** | 28.2 (21.0 - 28) | 45.5 (22.5 - 60) | ** |
| **NEWS** | 2.56 (1 - 4) | 7.27 (5 - 10) | *** | 3.47 (1 - 6.0) | 7.40 (6 - 10.8) | *** |
| **Hypertension** | 28 (39%) | 26 (53%) |  | 35 (39%) | 19 (61%) | * |
| **Cardiac disease** | 20 (28%) | 17 (35%) |  | 25 (28%) | 12 (39%) |  |
| **Kidney disease** | 24 (34%) | 19 (39%) |  | 28 (31%) | 15 (48%) |  |
| **eGFR** | 67.7 (49 - 90) | 56.5 (30 - 90) | * | 67.7 (48.0 - 90.0) | 49.9 (26.5 - 68.5) | ** |
| **Liver disease** | 7 ( 9.9%) | 6 (12.2%) |  | 9 (10%) | 4 (13%) |  |
| **Malignancy** | 13 (18%) | 6 (12%) |  | 13 (15%) | 6 (19%) |  |
| **Fever** | 34 (48%) | 30 (61%) |  | 49 (55%) | 15 (48%) |  |
| **Cough** | 33 (46%) | 29 (59%) |  | 43 (48%) | 19 (61%) |  |
| **SOB** | 35 (49%) | 29 (59%) |  | 48 (54%) | 16 (52%) |  |
| **Pulse (BPM)** | 87.9 (72.0 - 102) | 102.8 (91.8 - 114) | *** | 90.6 (74 - 106) | 103.8 (92 - 114) | ** |
| **Temp. (°C)** | 37.1 (36.6 - 37.4) | 37.5 (36.7 - 38.1) | * | 37.2 (36.6 - 37.5) | 37.3 (36.7 - 37.9) |  |
| **BP (mmHg)** | 137 (120 - 151) | 123 (102 - 138) | ** | 135 (120 - 149) | 120 (102 - 130) | ** |
| **Respiratory rate** | 20.1 (17 - 22) | 26.6 (20 - 30) | *** | 21.7 (17.0 - 22) | 25.7 (19.2 - 30) | * |
| **Hb (g/L)**<br>male [133-168]<br>female [118-148] | 126 (111 - 141) | 124 (107 - 146) |  | 127 (113 - 140) | 120 ( 96 - 146) |  |
| **WBC (x10<sup>9</sup>/L)**<br>[3.5-11.0] | 8.28 (4.8 - 10.9) | 9.98 (5.9 - 11.7) | . | 8.63 (5.1 - 11.0) | 9.97 (6.5 - 11.6) |  |
| **Lymphs (x10<sup>9</sup>/L)**<br>[1.0-3.5] | 1.25 (0.75 - 1.4) | 1.06 (0.60 - 1.6) |  | 1.239 (0.7 - 1.50) | 0.994 (0.5 - 1.35) | . |
| **PLTS (x10<sup>9</sup>/L)**<br>[150-400] | 256 (176 - 306) | 236 (166 - 292) |  | 255 (175 - 305) | 226 (169 - 259) |  |
| **HCT (%)** | 0.372 (0.337 - 0.417) | 0.371 (0.329 - 0.436) |  | 0.375 (0.341 - 0.416) | 0.361 (0.287 - 0.443) |  |
| **ALT (U/L)**<br>male [≤41]<br>female [≤33] | 32.8 (13 - 31.8) | 47.1 (16 - 39.0) |  | 36.1 (15 - 39.0) | 46.6 (13 - 31.5) |  |
| **Urea (mmol/L)**<br>[2.5-7.8] | 8.29 (4.6 - 8.75) | 13.05 (5.2 - 19.00) | ** | 8.53 (4.4 - 8.8) | 15.12 (8.0 - 22.2) | *** |
| **Creatinine (μmol/L)**<br>male [59-104]<br>female [45-84] | 112 (66 - 104) | 133 (70 - 168) |  | 109 (65.0 - 105) | 153 (84.5 - 184) | . |
| **CRP (mg/L)**<br>[<4] | 47.7 (11 - 61.2) | 132.9 (49 - 190.0) | *** | 61.4 (13.5 - 79.5) | 144.3 (52.5 - 209.5) | *** |

### Sample preparation for metabolomics analysis

Patient serum samples were thawed at room temperature and maintained on ice throughout the sample preparation process. Samples were prepared by addition of 100 µL sample to a 2 mL Eppendorf containing 350 µL methanol (LC-MS grade) previously cooled at −80 °C and maintained on dry ice whenever possible. The mixture of serum and methanol was vortexed vigorously followed by centrifugation at 18,000 x *g* for 15 min at 4 °C to pellet proteins. Multiple 75 µL aliquots (for extraction replicates) of the resulting supernatant dried in a vacuum centrifuge (ScanVac MaxiVac Beta Vacuum Concentrator system, LaboGene ApS, Denmark) with no temperature application and stored at −80 °C until required for LC-MS/MS analysis.

Batch quality controls (QC) and conditioning QC samples were also prepared in this way by pooling serum samples for each batch. Inter-batch quality assurance (IQA) samples were prepared with the same protocol using pooled serum samples provided by RLUH. Therefore, those samples are not representative of the study samples, but representative of the general hospital population and sample acquisition and storage practices.

Additional inter-batch quality assurance samples were prepared using commercial pooled human serum (BioIVT, Lot BRH1413770, Cat: HMSRM, mixed gender 0.1 um filtered) spiked with internal standards. Here, 100 µL sample were added to a 2 mL Eppendorf containing 330 µL methanol (LC-MS grade) and 20 µL of internal standards mix (ISTDs) as described in (Muelas et al., 2020). The mixture of methanol and internal standards (ISTDs) was previously cooled at −80 °C and maintained on dry ice when adding serum.

Extraction blanks were prepared in the same way as serum samples replacing serum and ISTDs mix with 120 µL of water (LC-MS grade). Prior to analysis, samples were resuspended in 40 µL water (LC-MS), centrifuged at 17,000 x *g* for 15 min at 4 °C to remove any particulates and transferred to glass sample vials.

### LC-MS/MS analysis of patient serum samples

Untargeted LC-MS/MS data acquisition was performed as previously described (Muelas et al., 2020) and using published methodologies and guidelines (Broadhurst et al., 2018; Broadhurst and Kell, 2006; Brown et al., 2005; Dunn et al., 2011; Mullard et al., 2015). Data were acquired using a ThermoFisher Scientific Vanquish UHPLC system coupled to a ThermoFisher Scientific Q-Exactive mass spectrometer (ThermoFisher Scientific, UK). Samples for the longitudinal aspect of this study were analysed in the same way but using a ThermoFisher Scientific ID-X Tribrid mass spectrometer (ThermoFisher Scientific, UK). Mass spectrometer operation and details are provided in Supplementary Information.

Samples were analysed following guidelines set out in (Dunn et al., 2011) and (Broadhurst et al., 2018). Briefly, blank extraction samples were injected at the beginning and end of each batch to assess carry over and lack of contamination. QC samples, prepared by pooling equal aliquots of analytical samples in each batch, were applied to condition the analytical platform, enable reproducibility measurements and to correct for systematic errors within batches. Quality Assurance (QA) samples were also incorporated in every batch at regular intervals up to 12 samples per run. Two pools of hospital patient serum (referred as Inter-batch quality assurance (IQA)) and commercial serum (referred as SQA) were prepared at the beginning of the study and used in every batch allowing batch alignment in the data processing step. Isotopically labelled internal standards were added to SQA samples, prepared as previously described (Muelas et al., 2020), to monitor mass accuracy. IQA samples were subsequently used to correct across all batches. Four samples per batch were run with replicates to ensure reproducibility.

### LC-MS/MS data pre-processing and analysis

Untargeted compound pre-processing was used for the discovery study and a targeted area extraction was employed in the validation study. However, data acquisition in both studies was performed with the same untargeted LC-MS method.

#### Discovery study data pre-processing

Raw instrument data from all batches in .RAW file format were exported to Thermo Fisher scientific Compound Discoverer 3.1 (CD3.1) for deconvolution, alignment and annotation (full workflow and settings as described in (Muelas et al., 2020)) based on IQA samples.

Compound grouping was performed in CD3.1 where 6971 compounds in ESI+ and 3122 compounds in ESI-were retained. Retained compounds are selected based on presence in more than 80% of the IQA, CV less than 30% and signal 5 times higher than the blank injections. From those compounds 267 in ESI + and 142 in ESI-had identification in mzCloud with score higher than 70% and full match on Predicted Composition. For all data acquired, annotation and identification criteria were according to (Schymanski et al., 2014) and (Sumner et al., 2007).

#### Validation and longitudinal study data pre-processing

Areas for the 20 compounds previously selected for the model were extracted in Thermo Fisher Scientific Trace Finder software from the .RAW files. Compound fragmentation database was obtained from Compound Discoverer on previous runs. Quan master method was employed allowing to sum all events in case of compounds eluting in multiple peaks such as cytosine and cytosine-containing compounds where source fragmentation was observed, or butrylcarnitine where multiple elution peaks were observed. ICIS detection algorithm was employed for all compounds; however, other detection algorithm settings (e.g., peak detection strategy, peak threshold type) and retention times settings (i.e., detection type and RT window) were tuned individually to each compound allowing for controlled selection of the area in every sample. Exported areas were further processed in R for normalization and statistical analysis.

#### Area normalization and batch correction

A custom 2-step normalization was used to correct for small within-batch runtime drift and larger between-batch variations in both studies. To perform this, non-normalized peak areas from CD3.1 were exported as a .csv file. QC-based correction was performed in R (version 4.0.2) as discussed in (Dunn et al., 2011) using *fANCOVA* package. The QC correction was performed on each batch independently to remove runtime drift intensity variations. Subsequently, variation between batches was corrected in refence to the IQA mean of all batches. PCA plots comparing the results for different normalization approaches are included in the supplementary information (Figure S6).

#### Metadata and multiblock analysis

The following variables in the metadata were used for multivariate analysis: gender, age, BMI, Glasgow coma scale (GCS), NEWS, pulse, temperature, blood pressure, respiratory rate, Hb, WBC, Lymphs, PLTs, HCT, ALT, total bilirubin, urea, creatinine, eGFR, CRP, FiO_2_ (%) and O_2_ saturation (%). The description of the biochemical tests is presented in Table 4. There were 1.44% missing values in this meta data set and they were imputed by using *K*-NN imputation algorithm (Troyanskaya et al., 2001).

The multiblock analysis was performed on three data blocks: LC-MS ESI+/ESI- data and metadata. For each block, the data matrix was auto-scaled so that each variable has a mean of 0 and a standard deviation of 1. In addition, a block scaling factor which is the inverse of the square root of the number of variables of this block was applied to compensate differences in variance due the difference in the number of variables. A multiblock PCA (MB-PCA) model called consensus PCA (Smilde et al., 2003; Xu and Goodacre, 2012) was applied to the three blocks of data. The results of this MB-PCA model are consisted of one super scores matrix which represents the common trend of all the blocks and three block scores matrix which represents the pattern of each block under perspective of the common trend.

#### Selection of significant metabolic features

Background compounds and compounds detected in less than 25% of the samples were excluded from further analysis. Compounds were further filtered down based on False Discovery Rate (FDR) corrected *p*-value (i.e., *q*-value) significance < 0.05 and log_2_ fold change > 0.5 in severity (i.e., severe vs mild cases) and outcome (i.e., diseased vs discharged patients). *p*-values were calculated using the Mann-Whitney as the data did not satisfy normality assumption required for T-test. The usually adopted log transformation approach to satisfy T-test requirements also tended to overemphasize low abundance compounds.

Significant differences (q-value < 0.05 and absolute log_2_ fold change > 0.5) were found for 1143/601 compounds in ESI+/- when comparing severe against mild cases, with 1650/680 compounds (ESI+/-) when comparing between outcomes. For simplicity, due to the overlap in patients across the two comparisons performed (severity and outcome), the union of significant compounds across these two comparisons was taken forward for further analysis, with a total of 1987/973 ESI+/- compounds.

### Signal curation

Significant compounds were manually curated in CD3.1 based on the LC signal quality and MS spectra. LC quality was assessed based on batch retention time (RT) overlap and clear peak separation. MS spectra was evaluated on the detection of preferred ion i.e., [M+H]^+^ and [M-H]^-^ with at least 2 isotopes. Additionally, compounds where the signal was filled by CD gap fill option for more than 20% of the QC and 90% of the samples were also excluded.

Where compounds of interest were detected in both ESI+ and ESI-, the clearest signal was retained for further analysis. When necessary standards were run to confirm MS/MS, RT and signal intensity by polarity. Specifically, in the case of uridine and pseudouridine best separation and signal intensity detection of standards was achieved in ESI-, therefore negative polarity data was used for those compounds.

#### Pathway enrichment analysis

Pathway enrichment analysis was performed in MUMMICHOG (Li et al., 2013) version 2 incorporated in MetaboAnalyst (Pang et al., 2020). To match the MUMMICHOG requested *m/z* compound format, resolved masses retrieved form CD analysis were altered to achieve [M+H]^+^ adduct for ESI+ and ESI-results. MUMMICHOG adduct option was set to recognize exclusively [M+H]^+^ adducts. This allowed processing of ESI+ and ESI-results together and minimize false hits due to multiple adduct matches. Pathways with a high number of significant hits were further manually investigated and hits subsequently verified by MS, RT, MS/MS and match against standards when available.

#### Compound identity validation

Compounds retained for the model identity were validated against standards whenever possible. Standard validation was performed against RT and MS2 profile of 1 µM and 10 µM standard in water or appropriate solution. In some cases, e.g., cytosine derivatives, asparagine and ureidopropionate the standards were also spiked in to pooled study serum at 5 µM to validate the RT in the matrix.

MS2 profile exploration was performed in the case of ureidopropionate due to a poor MS2 capture in the untargeted data acquisition. A targeted Parallel Reaction Monitoring (PRM) method focused on the 133.06077 *m/z* ion at a limited scan range 66.7 to 200 m/z was performed. The quadrupole isolation window was set to 1.2 m/z reflecting the original method value. This setup allowed for the acquisition of the MS2 profile of this compound at any LC gradient stage for all ions in a 1.2 m/z window range around 133.06077 m/z. This approach provided best results in capturing low intensity signal in areas of the LC gradient with high event load as was the case with ureidopropionate.

In the case of ‘180.05336@4.775‘, which was initially identified as nicotinuric acid at MSI level 3, further validation demonstrated that despite significant fragmentation profile overlap with nicotinuric acid the compound of interest structure is undeniably different based on LC gradient elution. Therefore, its identity remains unknown. More details related to compound identity validation and area calculation choices are available in the Supplementary information Sup. Section 1.1.

#### Multiple predictor models

Reported results for multiple predictor models were produced with a Bayesian logistic regression model implemented in R with *rstanarm* R package (Goodrich, 2020). Comparative analysis was performed with the extreme gradient boosting *xgboost* R package (Chen and Guestrin, 2016; Friedman, 2001), Logistic regression with Generalized Linear Models (GLM) *glm* R package and GLM with Elastic net regularization *glmnet* R package (Zou and Hastie, 2005). Data were separated into training and test groups (80:20) with balanced label ratios. Conservative regularization parameters were used to reduce overfitting. Bayesian GLM approach was set to increased regularization with prior scale = 1, for glmnet *alpha* was set to 0.1 and xgboost *eta* to 0.01 with *max_depth* = 1. Due to the large group disparities in outcome, weights were incorporated in the model to handle class imbalance. Sparsity inducing parameters such as L1 regularization in glmnet and lasso or hierarchical shrinkage prior in Bayesian GLM were avoided despite better results in some cases. This allowed to perform subgroup selection accounting for compound identity confidence level and known biological role. From the four compared methods Bayesian logistic regression consistently showed better generalization and was therefore retained for results reporting.

Mean and standard deviation for accuracy and AUC were estimated using cross-validation with 100 iterations. These cross-validation results were used to describe the model sensitivity to the data and not for hyperparameters optimization. Visual ROC and 95 % CI representations were obtained from a randomly selected train/test partition using pROC R package with 2000 stratified bootstrap replicates on the test data.

#### Adjusted compounds significance

Individual OR and 95% CI for selected compounds were evaluated with univariate logistic regression with Generalized Linear Models (*glm*) implementation in R *stats* package. OR (95% CI) and *p*-values for significance are presented in the reported tables. Compounds are adjusted for age, gender, BMI, liver disease, cardiovascular disease, hypertension, kidney disease (i.e., chronic kidney disease stages 2 to 5), diabetes mellitus (type 1 or 2) and all together. Liver conditions include cirrhosis, hepatitis, alcoholic hepititis, autoimmune hepatitis, ascites, transplant, fatty liver disease. Cardiovascular conditons include ischaemic heart disease (IHD), atrial fibrillation (AF), cardiomegaly, cardiomyopathy, left ventricular systolic dysfunction, left ventricular hypertrophy, left ventricular failure, congestive heart failure, drug induced myocarditis, heart failure, angina and Takotsubo cardiomyopathy with IHD and AF being most frequent.

## Data Availability

MTBLS2997 www.ebi.ac.uk/metabolights/

https://www.ebi.ac.uk/metabolights/

## Acknowledgements

We thank the UK BBSRC (grant BB/V003976/1) and the Novo Nordisk Foundation (grant NNF20CC0035580) for financial support.

## Contributions

D.B.K. and R.G. conceived the study and obtained funding for a grant to enable this work.

A.S.D and J.M.T. obtained ethical approval, acquired samples and metadata.

A.S. created a data base to store data.

M.W.M. and I.R. designed LC-MS/MS acquisition with advice from Y.X., D.B.K. and R. G.

M.W.M., J.M.G., N.G. and I.R. prepared samples and performed LC-MS/MS acquisition

M.W.M and I.R. processed data. I.R. performed data analysis, with Multi-block analysis performed by Y.X.

M.W.M. and I.R. performed data interpretation and wrote the manuscript with D.B.K.

ALL authors read and approved the final version of the manuscript.

## Data and code availability

LC-MS data for the discovery and validation studies in mzML format is freely available at MetaboLights repository (Haug et al., 2019) with study identifier MTBLS2997 (www.ebi.ac.uk/metabolights/).

The code covering data normalization, predictive models and basic visualizations is available at https://github.com/dbkgroup/COVID.

## Abbreviations

AEX-LC-MS/MS: anion-exchange LC-MS
AF: atrial fibrillation
ALT: alanine aminotransferase
ARDS: acute respiratory distress syndrome
AUC: area under the curve
BMI: body mass index
BP: systolic blood pressure
CI: confidence interval
COVID-19: Coronavirus disease 2019
CPAP: continuous positive airw ay pressure
CRP: C-reactive protein
CRS: cytokine release storms
CV: coefficient of variation
eGFR: estimated glomerular filtration rate
ESI: electrospray ionization
FC: fold change
FDR: false discover
FIO_2_: fraction of inspired oxygen
GC-MS: gas hromatography mass spectrometry
GCS: Glasgow coma scale
GLM: generalized inear models
Hb: haemoglobin evels
HCT: haematocrit
IHD: ischaemic heart disease
IQA: inter-batch quality assurance
LC-MS: liquid chromatography mass spectrometry
LC-MS/MS: liquid chromatography tandem mass spectrometry
LFC: log fold change
Lymphs: lymphocyte count
MB-PCA: multiblock PCA
MSI: Metabolomics Standards nitiative
MSML: mass spectrometry metabolite library
NEWS: national early warning score
NICE: national institute for health and care excellence
NMR: nuclear magnetic resonance
OR: odds ratio
PCA: principal component analysis
PCR: polymerase chain reaction
PLTs: platelets count
QC: quality control
RLUH: royal Liverpool university hospital
ROC: receiver operating characteristic
rRNA: ribosomal RNA
RT: retention time
RTPCR: reverse transcription PCR
SARS-CoV-2: severe acute respiratory syndrome coronavirus-2
SD: standard deviation
SOB: shortness of breath
WBC: white blood cell count

