## Supplementary information for "Untargeted metabolomics of COVID-19 patient serum reveals potential prognostic markers of both severity and outcome"

#### 1 Supplementary results and figures

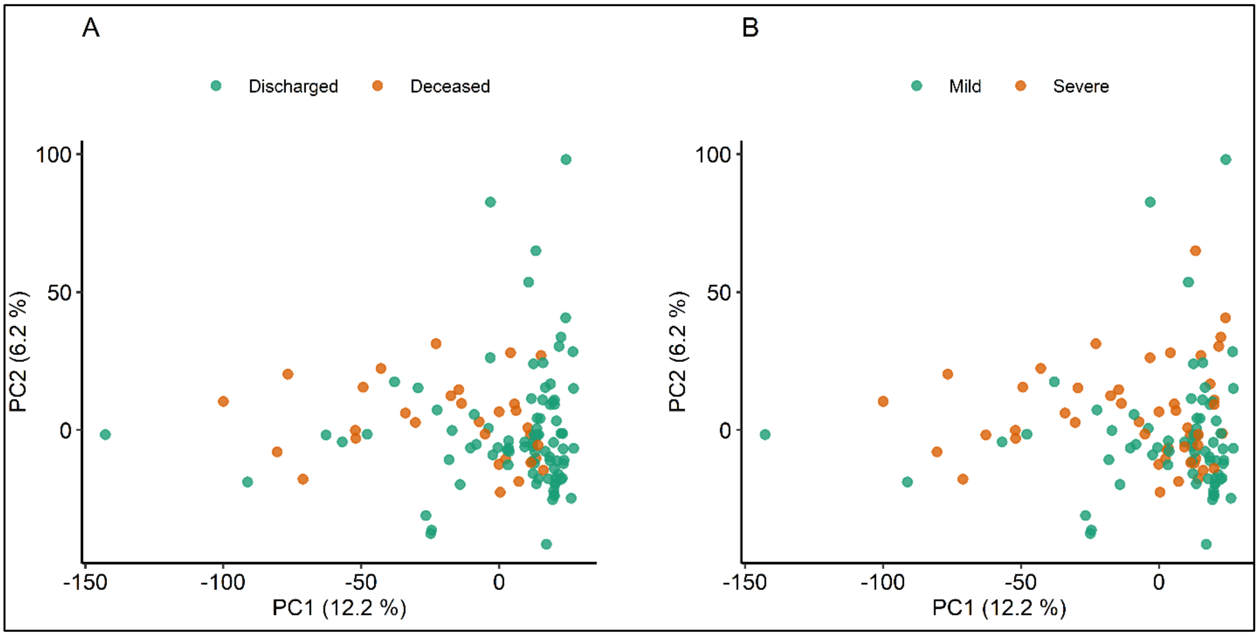


**Figure S1: PCA scores plot of the 120 patients coloured by outcome(A) and severity(B).** It can be observed that no clear group separation is visible. However, mild cases and discharged patients tent to cluster in the bottom right corner. Moreover, we can note that only relatively small percentage of the variance is explained by the first two components. Axis are labelled with principal component and its explained variance in percentage.

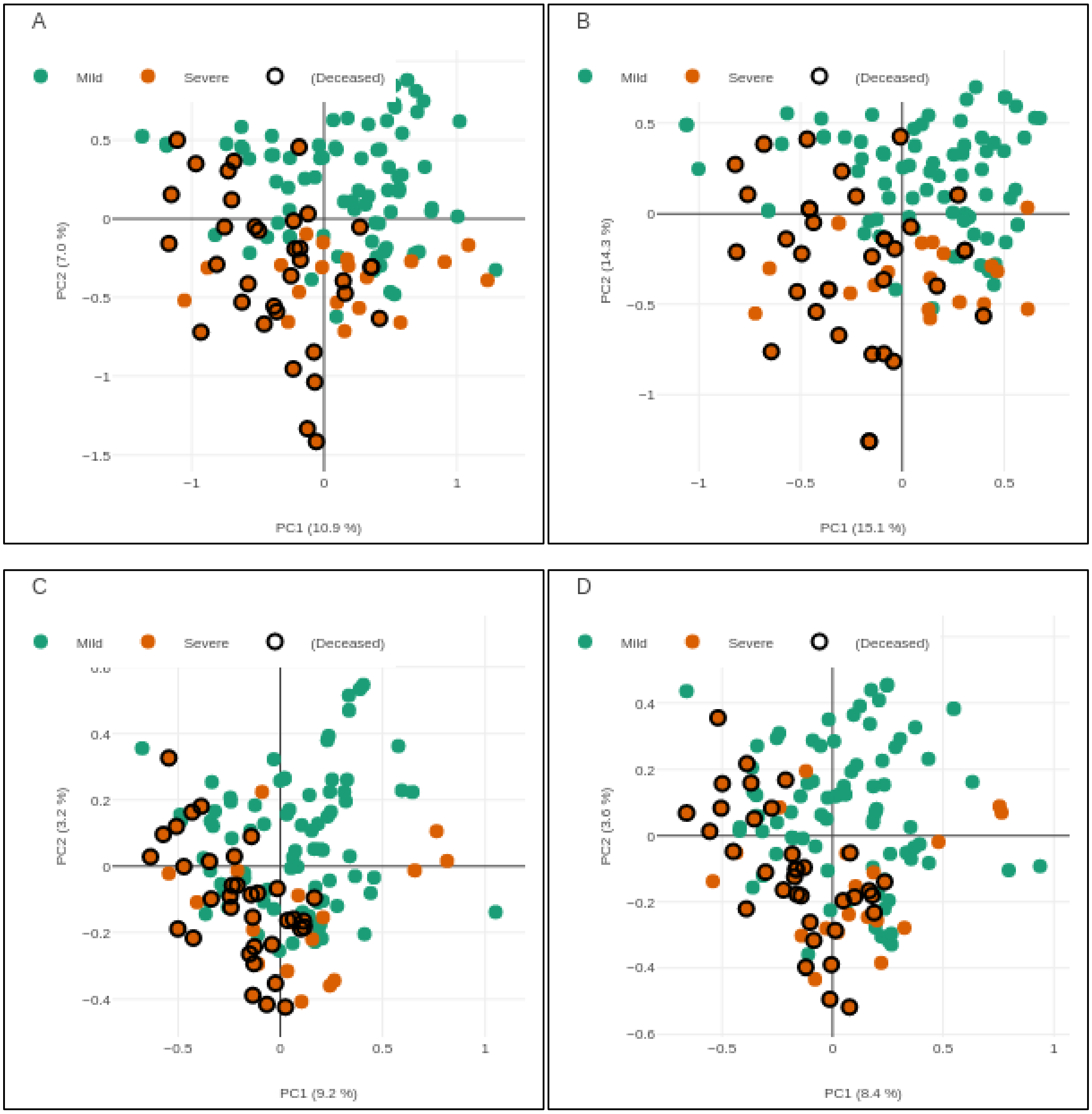


**Figure S2: Multiblock PCA in severity and outcome.** Super scores plot (A), followed by individuals blocks scores with metadata (B) LC-MS ESI+ (C) and LC-MS ESI- (D). Axes are labelled with principal component and its explained variance in percentage.

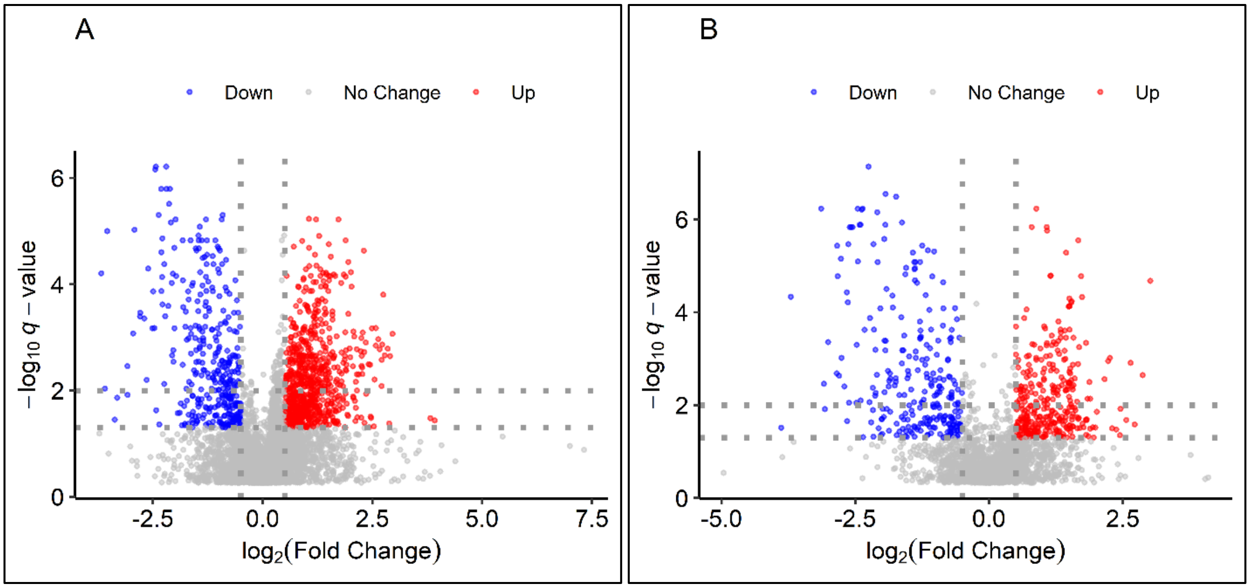


**Figure S3: Volcano plots ESI+ (A) and ESI- (B) for metabolites discriminating infection severity.** Dotted lines mark boundary of significance at $Log_2$ FC at 0.5 and *q*-value at 0.05 with second dotted line at *q*-value at 0.01. Compounds with higher levels in poor outcome are coloured in red and inversely compounds with significantly lower levels in blue.

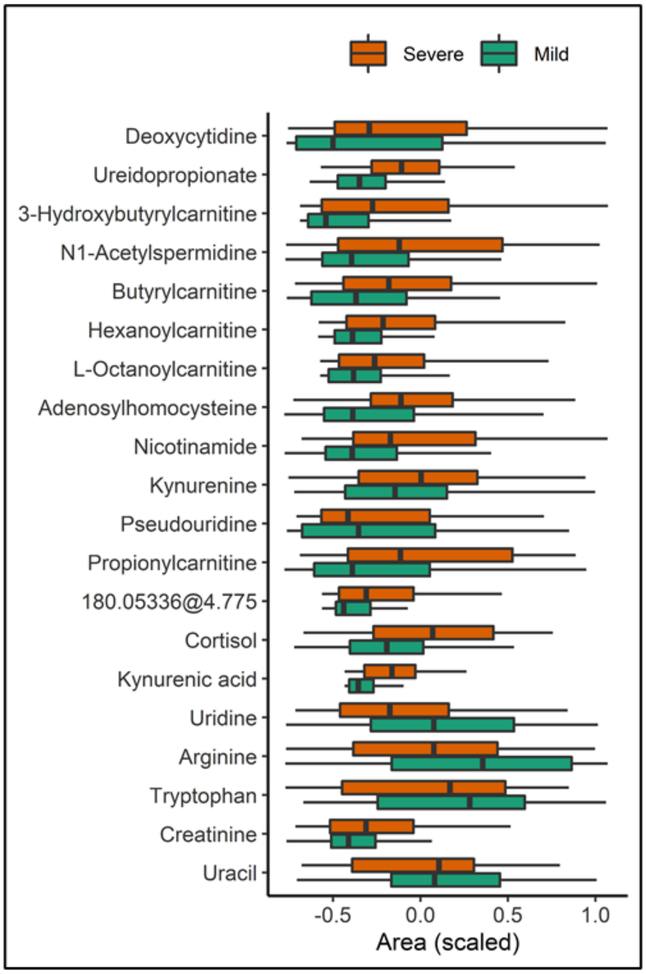


Compounds retained for severity and outcome predictive model and their significance in severe cases

| Name | Polarity | MSI | FC | *q*-value |
|---|---|---|---|---|
| Deoxycytidine | ESI+ | 1 | 3.70 | *** |
| Ureidopropionate | ESI+ | 1 | 3.30 | *** |
| 3-hydroxybutyrylcarnitine | ESI+ | 3 | 2.88 | ** |
| N1-acetylspermidine | ESI+ | 1 | 2.27 | *** |
| Butyrylcarnitine | ESI+ | 1 | 2.20 | * |
| Hexanoylcarnitine | ESI+ | 2 | 2.15 | ** |
| L-Octanoylcarnitine | ESI+ | 1 | 1.97 | * |
| Adenosylhomocysteine | ESI+ | 1 | 1.79 | ** |
| Nicotinamide | ESI+ | 1 | 1.59 | * |
| Kynurenine | ESI+ | 1 | 1.57 | *** |
| Pseudouridine | ESI- | 1 | 1.49 | . |
| Propionylcarnitine | ESI+ | 1 | 1.49 | * |
| '180.05336@4.775' | ESI+ | 3 | 1.46 | |
| Cortisol | ESI+ | 1 | 1.44 | ** |
| Kynurenic acid | ESI- | 1 | 1.29 | ** |
| Uridine | ESI- | 1 | 0.80 | ** |
| Arginine | ESI+ | 1 | 0.81 | * |
| Tryptophan | ESI+ | 1 | 0.84 | * |
| Creatinine | ESI+ | 2 | 1.13 | |
| Uracil | ESI+ | 1 | 0.93 | |

**Figure S4: Compounds retained for the predictive model and their significance in severity.** Box plot shows compound area differences between mild and severe COVID-19 patients ordered by fold change. Compounds areas are scaled by the mean to facilitate comparison. The table on the right side of the figure shows detailed information about the compounds including Fold Change (FC) and FDR corrected *p*-value (i.e. *q*-value) followign 'star' notation i.e. '***' correspond to *q*-values <0.001, '**' <0.01, '*' <0.05, '.' < 0.1 and missing when >0.1.

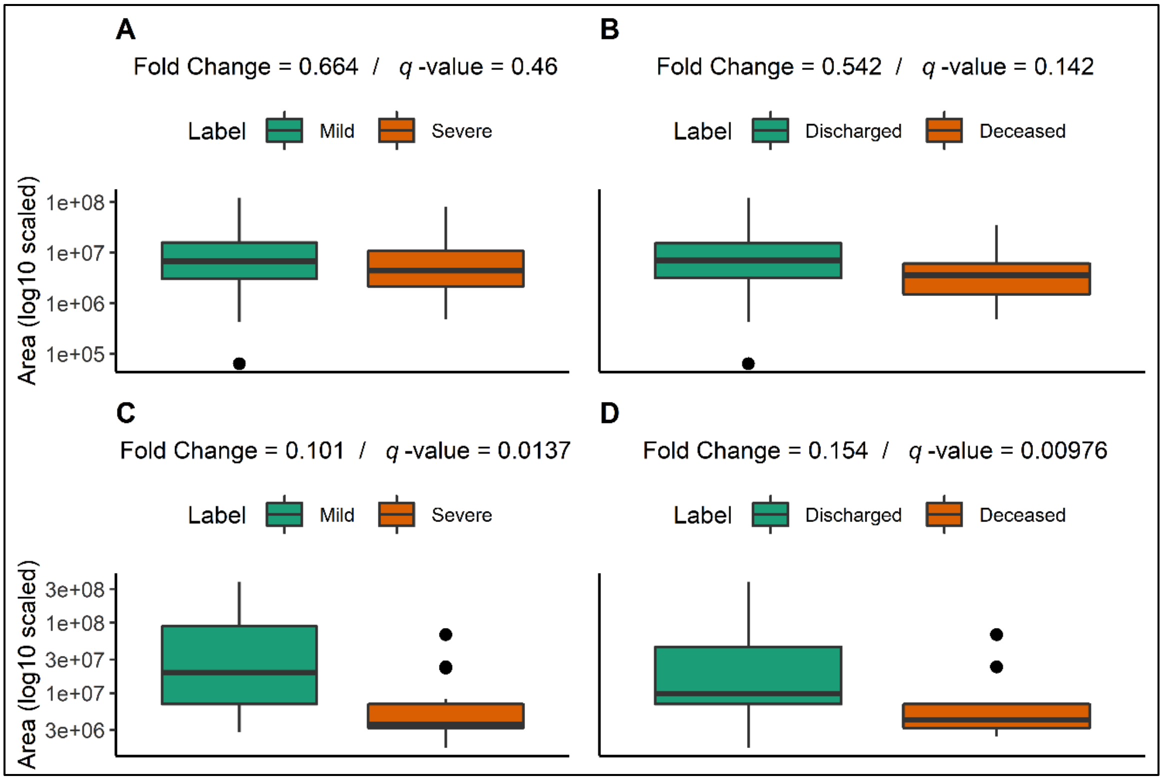


**Figure S5: Serum Ergothioneine in patients based on (A) severity and (B)outcome and piperine in severity (C) and outcome (D).** Ergothioneine shows expected trend (reduced in severe); however, falls short of q-value significance. The observed trend is stronger in poor outcome i.e. decreased levels of ergothioneine are more likely in poor outcome. Piperine levels appear significantly decreased in severe cases and poor outcome.

**Table S1: Adjusted logistic regression results by severity. Positive** OR indicate increased levels in patients with poor outcome. OR are presentred with OR(95%CI). Significance is presentedin 'star' notation i.e. '***' correspond to *p*-values <0.001, '**' <0.01, '*' <0.05, '.' < 0.1 and missing when >0.1.

| | Adjusted by: | None | | Age | | Gender | | BMI | | Liver disease | | CV disease | | Hypertension | | Kidney disease | | Diabetes | | All | |
|---|---|---|---|---|---|---|---|---|---|---|---|---|---|---|---|---|---|---|---|---|---|
| | Compound | OR | Sig. | OR | Sig. | OR | Sig. | OR | Sig. | OR | Sig. | OR | Sig. | OR | Sig. | OR | Sig. | OR | Sig. | OR | Sig. |
| Nucleic acids metabolism | Deoxycytidine | 6.3 (2.7-18) | *** | 4.8 (2.3-11) | *** | 7.2 (3-22) | *** | 4.3 (2.1-9.6) | *** | 6.5 (2.8-19) | *** | 6.3 (2.7-18) | *** | 6.5 (2.7-19) | *** | 6.3 (2.7-18) | *** | 6.3 (2.7-18) | *** | 7.2 (2.9-22) | *** |
| | Uracil | 0.81 (0.52-1.2) | | 0.82 (0.53-1.2) | | 0.86 (0.55-1.3) | | 0.87 (0.55-1.3) | | 0.83 (0.53-1.3) | | 0.79 (0.51-1.2) | | 0.82 (0.53-1.2) | | 0.8 (0.51-1.2) | | 0.8 (0.51-1.2) | | 0.83 (0.5-1.3) | |
| | Ureidopropionate | 120 (13-1853) | *** | 13 (4-55) | *** | 109 (12-1717) | *** | 9.5 (2.9-39) | *** | 116 (13-1817) | *** | 126 (14-1984) | *** | 128 (15-1992) | *** | 152 (15-2720) | *** | 152 (15-2720) | *** | 57 (7.5-861) | *** |
| | Pseudouridine | 1.6 (1.1-2.7) | * | 1.7 (1-2.9) | * | 1.6 (1-2.7) | . | 1.4 (0.92-2.4) | | 1.6 (1.1-2.7) | * | 1.6 (1-2.8) | * | 1.6 (1-2.7) | . | 2.3 (1.3-4.7) | * | 2.3 (1.3-4.7) | * | 2.1 (1.1-4.5) | * |
| | Uridine | 0.51 (0.3-0.81) | ** | 0.51 (0.3-0.82) | ** | 0.54 (0.32-0.86) | * | 0.59 (0.34-0.94) | * | 0.52 (0.31-0.83) | ** | 0.5 (0.29-0.8) | ** | 0.51 (0.3-0.82) | ** | 0.47 (0.26-0.77) | ** | 0.47 (0.26-0.77) | ** | 0.51 (0.27-0.87) | * |
| Kynurenine metabolism | Tryptophan | 0.62 (0.39-0.95) | * | 0.62 (0.38-0.96) | * | 0.58 (0.36-0.91) | * | 0.63 (0.38-0.99) | . | 0.62 (0.39-0.95) | * | 0.62 (0.39-0.96) | * | 0.62 (0.39-0.95) | * | 0.6 (0.37-0.94) | * | 0.6 (0.37-0.94) | * | 0.51 (0.29-0.84) | * |
| | Kynurenine | 3.4 (1.8-7.4) | *** | 3.7 (1.9-8.5) | *** | 3.2 (1.7-6.9) | *** | 2.7 (1.5-5.3) | ** | 3.4 (1.8-7.6) | *** | 3.6 (1.9-8.1) | *** | 3.3 (1.8-7.5) | ** | 4.2 (2-10) | *** | 4.2 (2-10) | *** | 4.6 (2.1-12) | *** |
| | Kynurenic acid | 1.1 (0.74-2) | | 1.1 (0.72-1.9) | | 1.1 (0.71-1.9) | | 1 (0.62-1.6) | | 1.1 (0.71-2) | | 1.1 (0.74-2) | | 1.1 (0.7-1.9) | | 1.2 (0.73-2.2) | | 1.2 (0.73-2.2) | | 0.96 (0.57-1.8) | |
| | Cortisol | 1.7 (0.99-3.6) | | 1.6 (0.97-3.2) | | 1.7 (0.99-3.6) | | 1.4 (0.88-2.9) | | 1.8 (1-4) | . | 1.7 (0.99-3.6) | | 1.7 (0.99-3.6) | | 1.7 (0.99-3.7) | | 1.7 (0.99-3.7) | | 1.5 (0.86-3.5) | |
| | Nicotinamide | 1.9 (1.1-3.6) | * | 2 (1.2-4.1) | * | 1.9 (1.1-3.7) | * | 1.8 (1-3.5) | . | 1.8 (1.1-3.6) | . | 2 (1.1-4) | * | 1.9 (1.1-3.7) | * | 1.9 (1.1-3.7) | * | 1.9 (1.1-3.7) | * | 2 (1-4.6) | . |
| Fatty acyl carnitines | Propionylcarnitine | 1.8 (1.1-3.1) | * | 1.8 (1.1-3) | * | 1.7 (1.1-2.9) | * | 1.6 (0.99-2.6) | . | 1.8 (1.1-3.1) | * | 1.8 (1.1-3.1) | * | 1.8 (1.1-3) | * | 1.9 (1.2-3.2) | * | 1.9 (1.2-3.2) | * | 1.5 (0.93-2.7) | |
| | Butyrylcarnitine | 3.1 (1.6-7.7) | ** | 2.7 (1.5-5.6) | ** | 2.9 (1.5-7.2) | ** | 2.2 (1.2-4.7) | * | 3.2 (1.6-8) | ** | 3.1 (1.5-7.9) | ** | 3 (1.5-7.6) | ** | 3.3 (1.6-8.5) | ** | 3.3 (1.6-8.5) | ** | 2.6 (1.3-6.8) | * |
| | 3-hydroxybutyrylcarnitine | 2.6 (1.5-5.7) | ** | 2.5 (1.4-4.9) | ** | 2.6 (1.5-5.7) | ** | 2 (1.2-3.9) | * | 2.7 (1.5-5.9) | ** | 2.6 (1.4-5.7) | ** | 2.6 (1.4-5.8) | ** | 2.7 (1.5-6) | ** | 2.7 (1.5-6) | ** | 2.3 (1.3-5.2) | * |
| | Hexanoylcarnitine | 2.3 (1.3-4.9) | * | 2.2 (1.3-4.5) | * | 2.3 (1.3-4.8) | * | 1.8 (1-3.4) | . | 2.3 (1.3-4.9) | * | 2.3 (1.3-5) | * | 2.2 (1.3-4.7) | * | 2.4 (1.3-5.4) | * | 2.4 (1.3-5.4) | * | 1.9 (1-4.3) | . |
| | L-Octanoylcarnitine | 1.9 (1.1-3.4) | * | 1.9 (1.1-3.5) | * | 1.8 (1.1-3.4) | * | 1.5 (0.93-2.6) | | 1.9 (1.1-3.5) | * | 1.9 (1.1-3.4) | * | 1.8 (1.1-3.3) | * | 2 (1.2-3.7) | * | 2 (1.2-3.7) | * | 1.6 (0.91-3.1) | |
| Other | Creatinine | 1.1 (0.75-1.8) | | 1.1 (0.73-1.8) | | 1.1 (0.71-1.7) | | 1 (0.67-1.7) | | 1.1 (0.76-1.8) | | 1.2 (0.76-1.8) | | 1.1 (0.7-1.8) | | 1.2 (0.74-2.2) | | 1.2 (0.74-2.2) | | 1.1 (0.66-2) | |
| | Arginine | 0.5 (0.3-0.79) | ** | 0.5 (0.29-0.79) | ** | 0.5 (0.3-0.8) | ** | 0.6 (0.36-0.95) | * | 0.49 (0.29-0.78) | ** | 0.49 (0.29-0.78) | ** | 0.51 (0.3-0.81) | ** | 0.46 (0.27-0.75) | ** | 0.46 (0.27-0.75) | ** | 0.52 (0.29-0.87) | * |
| | N1-acetylspermidine | 5 (2.2-14) | *** | 3.7 (1.9-8.3) | *** | 4.9 (2.2-14) | *** | 3.1 (1.6-7) | ** | 5 (2.2-14) | *** | 5 (2.2-14) | *** | 5.2 (2.2-15) | *** | 5.7 (2.4-17) | *** | 5.7 (2.4-17) | *** | 5.2 (2.1-17) | ** |
| | '180.05336@4.775' | 1.3 (0.84-2.2) | | 1.3 (0.82-2.1) | | 1.2 (0.77-2.1) | | 1.2 (0.75-1.9) | | 1.3 (0.84-2.2) | | 1.3 (0.84-2.2) | | 1.2 (0.8-2.1) | | 1.4 (0.86-2.7) | | 1.4 (0.86-2.7) | | 1.3 (0.74-2.3) | |
| | Adenosylhomocysteine | 2.6 (1.3-5.8) | * | 2.4 (1.3-5) | * | 2.5 (1.3-5.7) | * | 2 (1.1-4.3) | * | 2.5 (1.3-5.6) | * | 2.6 (1.3-5.8) | * | 2.5 (1.3-5.6) | * | 3.6 (1.6-9.2) | ** | 3.6 (1.6-9.2) | ** | 2.8 (1.3-7.6) | * |

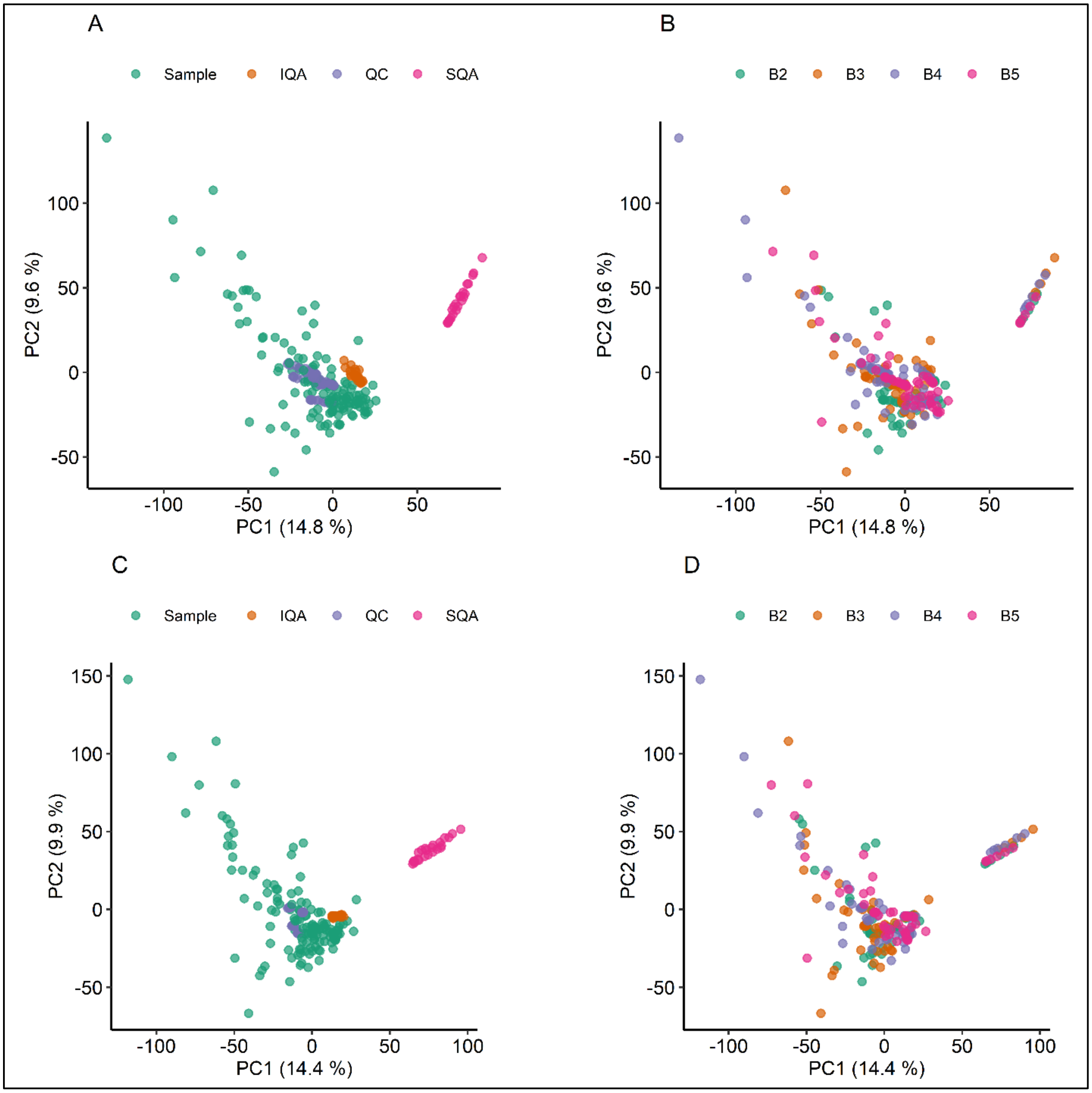


**Figure S6: PCA scores plot showing normalization results.** A and B shows raw data coloured by group and batch, respectively. C and D plots show normalized data. QC samples in plot C do not overlap between batches as expected because they represent a pool of the samples in each batch therefore are different between batches. SQA samples show reduced but still present drift after normalization. This is due to the different nature of those samples as not all compounds present in the commercial serum were measured in the study samples (e.g. recreational drugs). Finally, batches after normalization show a healthy distribution with no visible batch effect. Axis are labelled with principal component (PC) and its explained variance in percentage.

#### 1.1 Compound identity validation

Compound identity validation was performed when necessary to establish preferentially MSI 1 level for all selected compounds. Validation criteria were based on fragmentation spectra similarity (i.e., fragments and intensity levels) and overlap in RT in the LC method used. In cases more complex cases e.g., deoxycytidine and ureidopropionate, presented in detail in this section, pooled serum from the study was spiked with standards to establish clear RT overlap in matrix.

##### 1.1.1 Deoxycytidine

Cytidine and deoxycytidine fragment at the source resulting in cytosine events on the LC separated by approx. 0.1 min for cytosine, cytidine and deoxycytidine respectively as demonstrated in Figure S7. This indicated that the N-glycosidic bond between the cytosine and ribose moiety breaks easily. Therefore, we analysed the cytosine source fragmentation in all cytosine-based nucleoside, ribonucleotides and deoxyribonucleotides. All except CDP and CTP showed loss of cytosine however, only cytidine and deoxycytidine had strong signal in ESI+. Figure S8 shows pooled study serum spiked with 5 µM of the chemical standard that overlays perfectly with the 3 cytosine events detected in the study samples. Study serum cytosine levels were particularly low and undetected in some patients leading to a possible confusion of cytidine or deoxycytidine for a cytosine detection.

In the case of deoxycytidine approximately 2/3 of the molecules fragment at the source (see Figure S9) making the cytosine event a more reliable measurement for the deoxycytidine levels. Therefore, in the discovery study we used cytosine event areas at the deoxycytidine retention time for all reported results. In the validation study all three cytosine events i.e., cytosine, cytidine and deoxycytidine were summed up together and are referred as cytosine-based nucleosides. At this stage of the study this level of precision is sufficient and allows simpler and more accurately comparable measurements.

##### 1.1.2 Ureidopropionate

Ureidopropionate tends to fragment at the source leading to low levels of precursor ions and unreliable MS2 acquisition. Its elution time follows the isobaric asparagine by 0.2 min leading to possible confusion of the two compounds. A parallel reaction monitoring (PRM) method was used to capture the fragmentation profile to confirm its identity by MS2 spectra capture in addition to chemical standard based RT validation. The PRM obtained MS2 spectra showed signs of background noise ions that were filtered out based on mass defect and a coeluting ion in the serum. However, the two identifying fragments for ureidopropionate, 90.0550 and 115.0502, showed linear increase with injection volume: for volumes of of 1, 5, 10 µL.

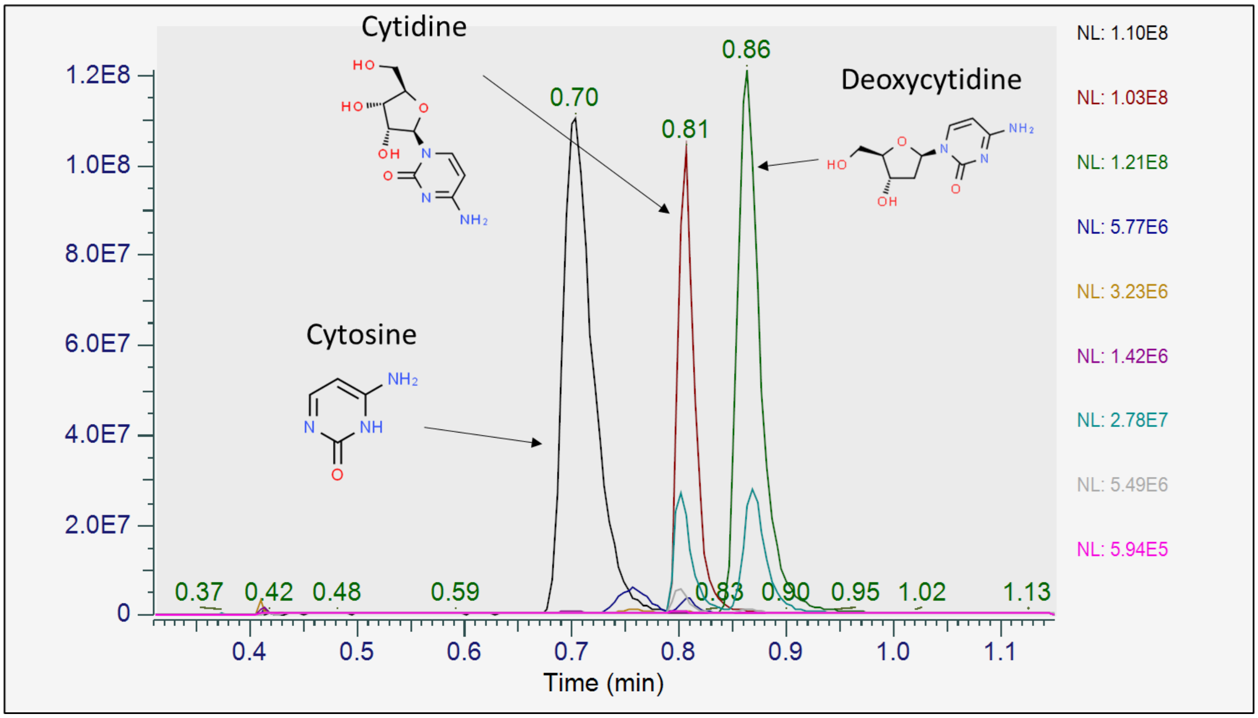


**Figure S7 Cytosine LC-MS ESI+ event from cytosine-based nucleoside, ribonucleotides and deoxyribonucleotides global scale overlay**. 1 µM of standard in water was acquired with the same method as the study separately and in a mix. All individual spectra in the figure are filtered for mass range event at 112.0505 m/z (cytosine). Cytosine identity of the source fragments was also confirmed by the corresponding MS2. All standards are overlayed in global range to allow for relative comparison. Cytosine (black), cytidine (red), deoxycytidine (green) shows the strongest signal indicating good ionization in ESI+. CMP (dark blue) and dCMP (cyan) are also captured relatively well. CDP (yellow), CTP (purple), dCDP (gray) and dCTP (magenta) are not noticeable in this overlay because they much lower intensity. In general ribonucleotides and their deoxy relatives are better detected in ESI-.

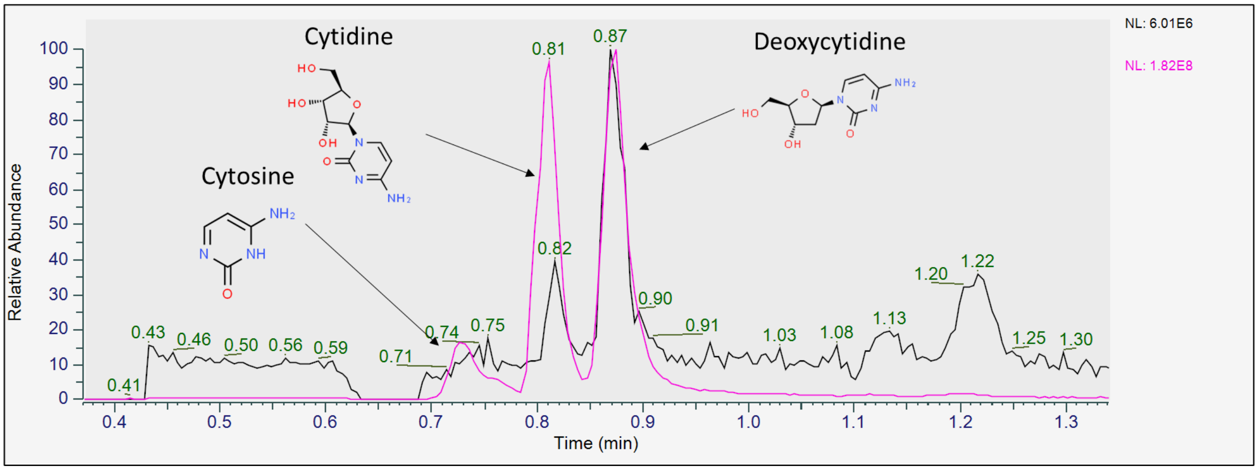


**Figure S8 Cytosine events in study serum with and without standards overlay in local scale.** Study pooled serum (black) and same serum with 5 µM cytosine-based nucleoside, ribonucleotides and deoxyribonucleotides standards (magenta) are overlayed in local scale. The three peak cytosine patterns clearly indicate cytosine at ~0.7min, cytidine at ~0.81 min and deoxycytidine at ~0.87 min. It is important to note that the serum signal is at 6.01e6 and the spiked serum signal at 1.82e8.

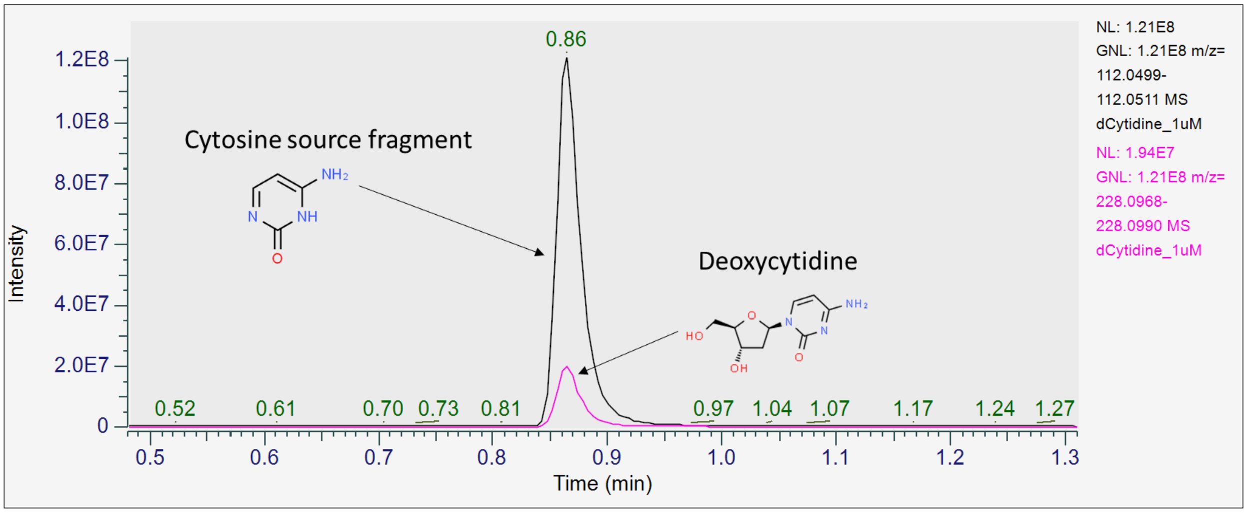


**Figure S9 Deoxycytidine fragmentation at the source.** Deoxycytidine 1µM in water filtered for cytosine event (black) and deoxycytidine event (magenta) at a global scale shows that the compound readily fragments at the source on the N-glycosidic bond between the cytosine and ribose moiety. Therefore, in the study we preferred to measure the cytosine event produced from the deoxycytidine. The overlap between both events in RT shows that the fragmentation is taking place post LC.

### 2 Supplementary methods

#### 2.1 MS acquisition settings for the analysis of longitudinal serum samples using an ID-X Tribrid mass spectrometer

As described in the main text, serum samples for longitudinal untargeted metabolomics were acquired in the same manner as baseline samples except a ThermoFisher Scientific ID-X Tribrid mass spectrometer was used. Instrument parameters are provided below.

Full-scan MS data was acquired in the Orbitrap mass analyser in the m/z range 66.7-1,000 with a mass resolution of 120,000 Full Width Half Maximum (FWHM) at m/z = 200, a chromatographic peak width to 4 s (FWHM), Normalized AGC target (%) = 50 and maximum injection time = 100 ms. Internal mass calibration was used in negative ionization mode only. Source and ion transfer parameters applied were as follows: Sheath flow rate (arbitrary units) = 40, auxiliary gas flow rate (arbitrary units) = 8, sweep gas flow rate (arbitrary units) = 1, spray voltage = 3.5 kV (positive) and -3.0 kV (negative), capillary temperature (°C) = 275, S-lens RF level (%) = 45, auxiliary gas heater temperature (°C) 300, source position = M 2.

Data-dependent MS/MS (ddMS$^2$) data acquisition was performed on pooled samples at the end of the analytical batch as described in guidelines by (Broadhurst *et al.*, 2018). In a manner similar to that proposed by (Mullard *et al.*, 2015), ddMS$^2$ data acquisition was performed on 4 precursor mass ranges in triplicate injections: 66.7-1,000, 66.7-300, 300-600 and 600-900. Data was acquired in the Orbitrap mass analyser with a mass resolution of 60,000 Full Width Half Maximum (FWHM) at m/z = 200 for the master scan and 30,000 FWHM at m/z 200 for ddMS$^2$ , both with a chromatographic peak width to 4 s (FWHM), Normalized AGC target (%) = 50, maximum injection time = 54 ms, cycle time (sec) = 0.6 , isolation window = 1.5 m/z, stepped HCD = 20, 40 and 60, AGC target Standard, intensity threshold = $2x10^4$, exclude isotopes = on and dynamic exclusion = 6.0 s.

The ThermoFisher Scientific AcquireX deep scan acquisition workflow was also employed here on pooled samples for each batch of longitudinal serum samples. This workflow enables an exhaustive acquisition of unique features in samples by using automated and iterative data dependent MS/MS acquisition, using automatically updated run-to-run inclusion and exclusion lists.
